# Abnormal T-Cell Activation And Cytotoxic T-Cell Frequency Discriminates Symptom Severity In Myalgic Encephalomyelitis/Chronic Fatigue Syndrome

**DOI:** 10.1101/2025.01.02.24319359

**Authors:** Ji-Sook Lee, Eliana Lacerda, Caroline Kingdon, Giada Susannini, Hazel M Dockrell, Luis Nacul, Jacqueline M Cliff

## Abstract

Myalgic encephalomyelitis/chronic fatigue syndrome (ME/CFS) is a debilitating but poorly-understood disease. ME/CFS symptoms can range from mild to severe, and include immune system effects alongside incapacitating fatigue and post-exertional disease exacerbation. In this study, we examined immunological profiles of people living with ME/CFS by flow cytometry, focusing on cytotoxic cells, to determine whether people with mild/moderate (n= 43) or severe ME/CFS (n=53) expressed different immunological markers. We found that people with mild/moderate ME/CFS had increased expression of cytotoxic effector molecules alongside enhanced proportions of early-immunosenescence cells, determined by the CD28^-^CD57^-^ phenotype, indicative of persistent viral infection. In contrast, people with severe ME/CFS had higher proportions of activated circulating lymphocytes, determined by CD69^+^ and CD38^+^ expression, and expressed more pro-inflammatory cytokines, including IFNγ, TNF and IL-17, following stimulation *in vitro*, indicative of prolonged non-specific inflammation. These changes were consistent across different cell types including CD8^+^ T cells, mucosal associated invariant T cells and Natural Killer cells, indicating generalised altered cytotoxic responses across the innate and adaptive immune system. These immunological differences likely reflect different disease pathogenesis mechanisms occurring in the two clinical groups, opening up opportunities for the development of prognostic markers and stratified treatments.

**Graphical Abstract.**
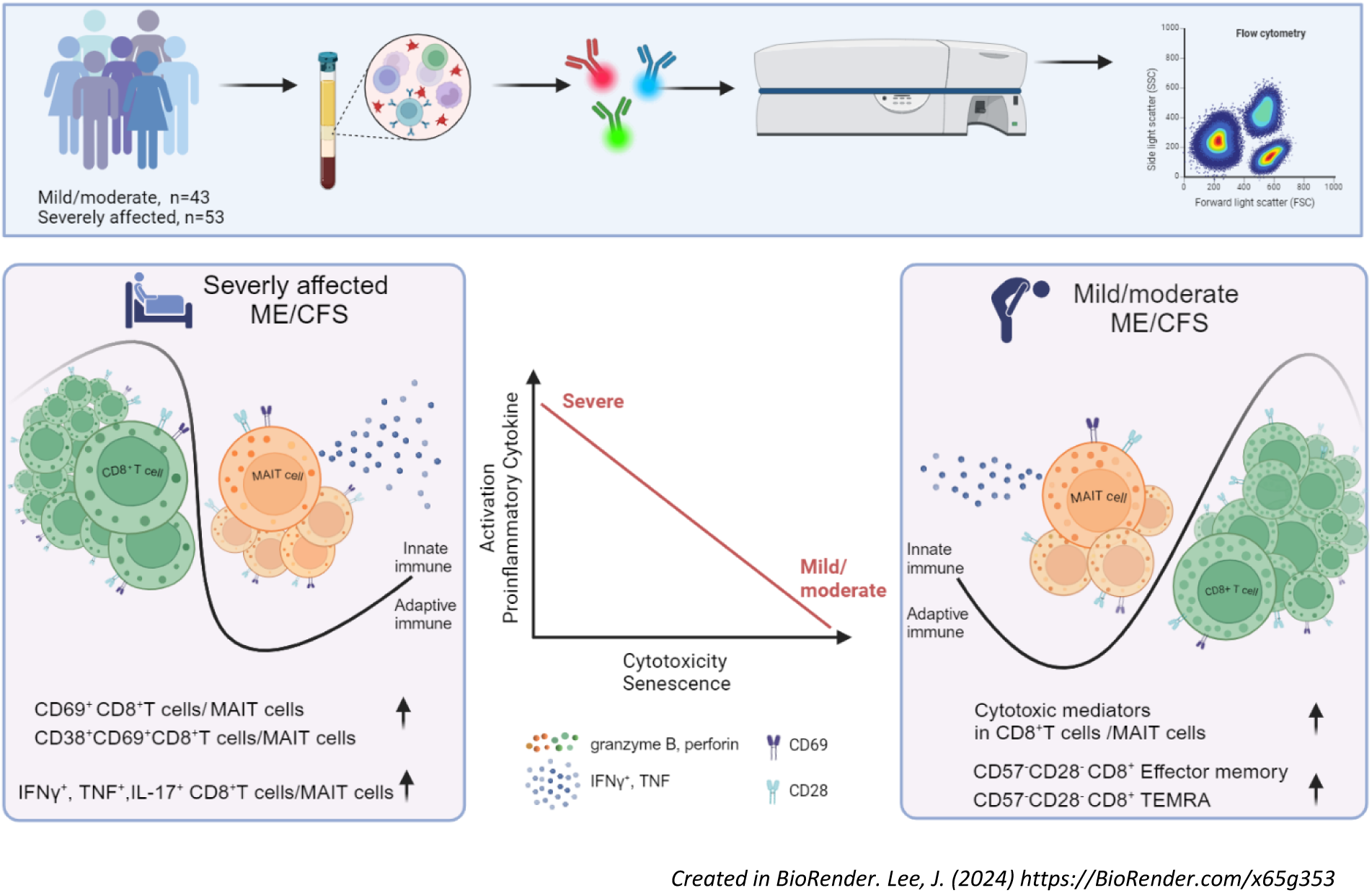

## Introduction

Myalgic encephalomyelitis/chronic fatigue syndrome (ME/CFS) is a debilitating disease with a prevalence of 0.1 to 2.2% in Europe (1). It is characterized by profound fatigue and post-exertional malaise (PEM), along with sleep, neurocognition, immune and autonomic nervous system dysfunctions, significantly impacting quality of daily life (2, 3). ME/CFS affects all age groups, including children and adolescents, and is more common in women than men (4). The symptom pattern is heterogeneous between individuals and can change within individuals through time (5); the majority of people living with ME/CFS (PWME) do not fully recover (6, 7). People with mild/moderate ME/CFS can often engage in some daily activities albeit with limitations, but those who are severely affected are usually housebound, and in some cases bedbound (8). Disease time-course patterns are diverse: more than half the PWME experience fluctuating (relapsing/remitting) symptom severity, particularly those with more mild/moderate disease; some have a gradual improvement in symptoms while others deteriorate long-term. As the pathophysiological mechanisms of this disease are still to be determined, diagnosis of ME/CFS is still based on clinical features (9), and the degree of disease severity has been classified by Carruthers *et al* into mild, moderate, severe, and very severe groups (10). The aetiology of ME/CFS is not fully understood; its onset often follows acute infections, but we know little about the triggering events for symptom fluctuation and the regulatory events which enable remission. We also do not know whether mild, moderate and severe ME/CFS are caused by the same disease mechanisms, or whether different symptom manifestations represent different underlying causal events. Better understanding of these differences would enable stratification of PWME, the development of personalised treatments and the possibility of elusive diagnostic biomarkers.

Immune system abnormalities have been investigated in PWME, to attempt to understand the disease aetiology. Altered T cell phenotype and activation, and impaired natural killer (NK) cell cytotoxicity have been reported (11–15), suggesting an increased susceptibility to infections, including reactivation or persistency of human herpesviruses (16, 17), or gut microbiome dysbiosis. However, recent reports showed no association or inconsistent results of the immune cell phenotype and function (18–21): most studies have analysed small cohorts, often with diverse clinical presentations, and clear identification of immune alterations in ME/CFS remains elusive. Overlapping symptom and laboratory results have been described between people with long COVID following SARS-CoV-2 infection and PWME, suggesting mechanistic similarity between the conditions (22, 23).

Clinical and immunological parameters have been shown to be associated with disease severity in ME/CFS. In clinical analyses, we previously reported handgrip strength as a potential diagnostic biomarker for ME/CFS (24), while blood creatine phosphokinase concentration was able to discriminate between people with mild/moderate and severe ME/CFS (25). Immunological studies have sought to determine serum cytokine signatures in ME/CFS associated with disease severity and disease duration. Montoya *et al* reported 13 inflammatory cytokines levels in serum were correlated with severity of disease (26), while people with recent onset of ME/CFS had prominent elevation of serum proinflammatory and anti-inflammatory cytokines which were not present in people with longer-term ME/CFS (27). Recently, natural regulatory autoantibodies were reported to be associated with symptom severity in people with ME/CFS which was triggered by infection (28). Further, abnormal increase of intracellular giant lipid organelles in peripheral immune cells has been reported in people severely affected with ME/CFS (29). From these studies it can be inferred that investigation of immunological parameters which distinguish between people with mild/moderate (ME-MM) or severe (ME-SA) ME/CFS will help our understanding of ME/CFS pathomechanisms. In our previous study, in a large cohort of ∼250 well-characterised PWME, we found alterations in the phenotype of CD8^+^ T cells, with an increase in effector memory cell and decrease in terminally differentiated cell proportions, alongside an elevated frequency of MAITs and of CD8^+^ MAITs, in people severely affected with ME/CFS (18): this study did not include a full analysis of cytotoxic cells, nor their functional capacity to produce cytokines or cytotoxic mediators such as perforin or granzymes.

Here, we aimed to determine how immunological parameters, particularly MAIT and CD8^+^ T cell phenotype and function, were associated with clinical symptom scores in PWME, to identify prognostic biomarkers and to facilitate better understanding of complex disease pathogenesis. To achieve this, we analysed samples from a cohort of people with ME-MM or ME-SA which were collected longitudinally (2 to 5 times at approximately 6-month intervals), and compared the flow cytometry data generated with eleven clinical parameters which were adapted for assessment of disease severity in PWME.

## Results

### Study Participants

The study population consisted of ninety-six individuals with ME/CFS, who were blood donors for the UK ME/CFS Biobank (30) (Table 1). All participants had a medically confirmed ME/CFS diagnosis from the UK National Health Service, and were rigorously assessed by the study clinicians to assure compliance with the Centers for Disease Control (CDC-94) (31) and/or Canadian Consensus Criteria (9). Participants were extensively clinically assessed, including measurement of creatinine phosphokinase in blood and handgrip strength reading by dynamometer (24). Clinical scores were calculated for each of the following domains based on the Canadian Consensus Criteria: post-exertional malaise, sleep dysfunction, pain, neurological/cognitive dysfunction, autonomic dysfunction, neurocognitive dysfunction and immune system manifestations. Forty-three (44.8%) of those research participants were classified at recruitment as having mild/moderate symptoms and were ambulatory (ME-MM), whereas 53 (55.2%) had severe symptoms and were house- or bed-bound (ME-SA).

**Table 1.** Demographic characteristics of study population.

| Variables |  | Mild/Moderate (n) | Severe (n) | P value |
| --- | --- | --- | --- | --- |
| Age |  | 38.5 | 40.1 | 0.328 |
| Sex | Male | 8 | 12 | 0.362 |
|  | Female | 35 | 41 |  |
| BMI | Male | 25.3 | 23.8 | 0.391 |
|  | Female | 24.5 | 24.8 |  |
| Handgrip strength |  | 20.8 | 18.1 | 0.180 |
| PCS |  | 30.3 | 19.4 | <0.001 |
| MCS |  | 42.3 | 45.0 | 0.039 |

**Note:** BMI – Body Mass Index, PCS – Physical Component Summary (from SF-36v<sup>2</sup><sub>TM</sub>), MCS – Mental Component Summary (from SF-36v<sup>2</sup><sub>TM</sub>).

Repeated blood samples were taken from each participant, to remove day-to-day fluctuations in cell number and function, and the average of these taken as the data point in the flow cytometric analysis. Participants were reassessed at each study visit: while there was some degree of fluctuation in disease severity at different timepoints, particularly within the ME-MM group, this was not of a sufficient degree to lead to changes in severity classification.

### Mucosal-associated invariant T cell frequency is elevated in ME-SA

*C*ryopreserved peripheral blood mononuclear cells (PBMCs) were thawed and analysed by flow cytometry: the gating strategy is shown in Supplemental Figure 1. Initially, we sought to confirm whether PWME had altered frequencies of T cell subsets and NK cells, depending on the severity of their symptoms, in this robust repeated-measures analysis. Firstly, we compared the frequencies of T cells (identified as CD3^+^) and T cell subsets (CD4^+^ T cells, CD8^+^ T cells, CD4^-^CD8^-^ Double Negative (DN) T cells, CD4^+^CD8^+^ Double Positive (DP) T cells, CD3^+^CD56^+^ NKT-like cells), as well as NK cells (identified as CD3^-^CD56^+^) and CD8^+^CD56^+^NK cells (“NK8 cells”) in *ex vivo* PBMCs from people with ME-MM or ME-SA, and found there were no differences between the two clinical groups (Supplemental Figure 2A-2H). In a previous study (18), we found that the frequency of circulating total MAIT cells (phenotype: CD3^+^TCR Vα7.2^+^CD161^++^) was elevated in ME-SA, compared to healthy controls, multiple sclerosis and ME-MM, and also that the frequency of CD8^+^ MAITs within total MAITs was significantly elevated in ME-SA compared to ME-MM and healthy controls. In this study, we included fluorescently labelled MHC related protein-1 (MR-1) tetramer staining for better identification of the MAIT subset within the CD3^+^ T cell population (32). As shown in Figure 1, we confirmed that the ME-SA group had elevated frequencies of CD3^+^ MAIT cells and of CD8^+^ MAITs, which is a major subset of total MAITs, compared to those in the ME-MM group. These patterns were reproducible through four different flow cytometry staining panels utilised, and the differences became more apparent after overnight resting in culture medium as part of the “*in vitro* stimulation” methodology. Interestingly, the frequency of CD4^+^ MAIT cells was higher in the ME-MM group (Figure 1C). There was no significant difference in CD4^+^CD8^+^ MAIT frequency between the two groups. We also confirmed that the frequencies of NK cells and the minor DN T cell subsets were reproducible across the different staining methods, including extracellular-only and additional intracellular staining: there was a slight increase in frequencies detected with the ‘function’ staining panel, but the pattern between the two clinical groups was highly similar across all staining methods (Supplemental Figure 2I). Interestingly, we observed significantly diminished expression of CD8, based on Median Fluorescence Intensity (MFI), on CD8^+^ T cells in the ME-MM group compared with the ME-SA group (Supplemental Figure 3A). This can be caused by the appearance of a CD8^low^ population: however, here there was no significant difference between ME-MM and ME-SA in the frequency of the CD8^low^ population (Supplemental Figure S3B).

**Figure 1.**
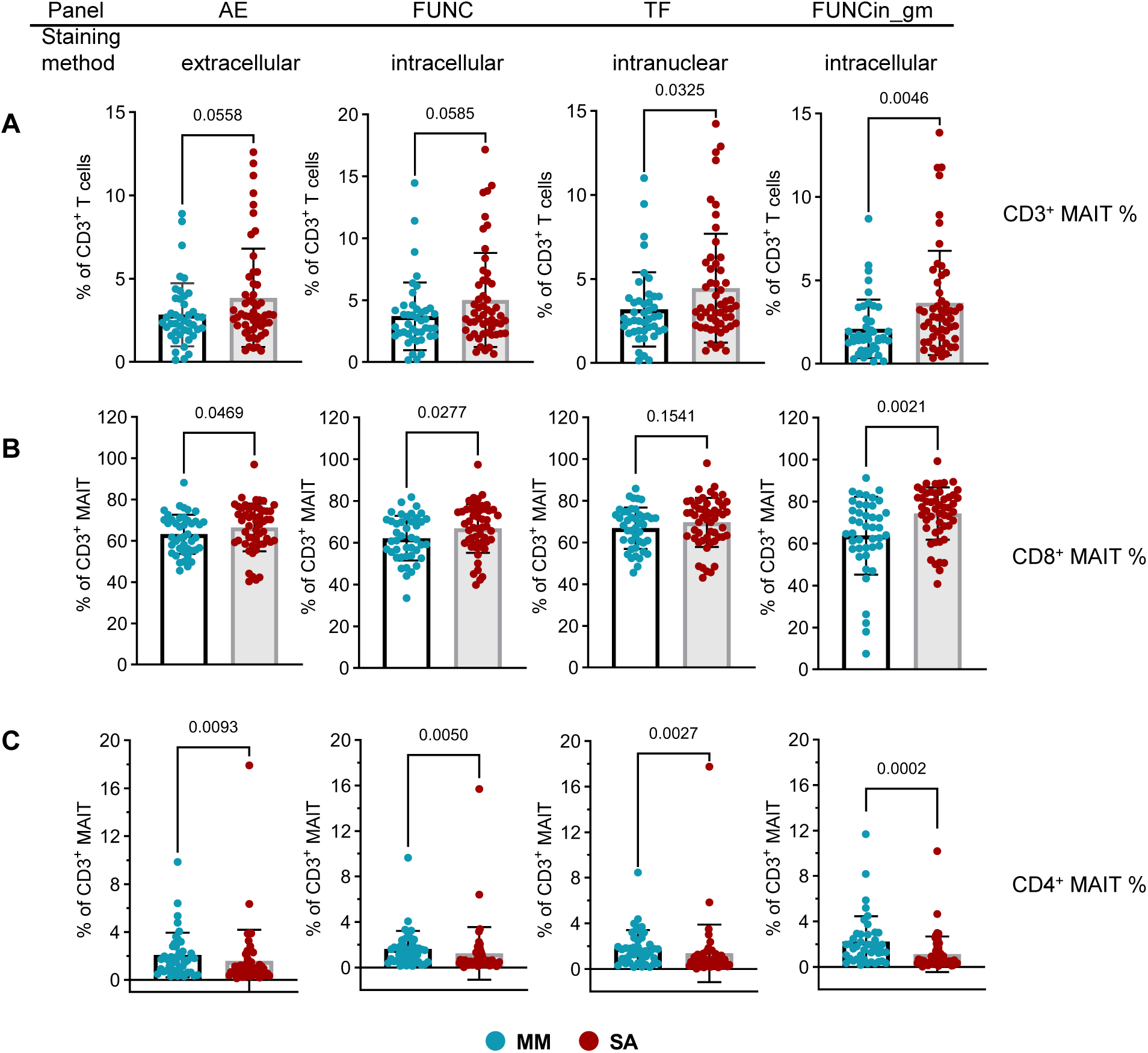
Differential frequencies of Mucosal-associated invariant T cell subsets in mild/moderate (n=43) and severe ME/CFS (n=53). PBMC from people with mild/moderate (MM) or severe (SA) ME/CFS symptoms were analysed by flow cytometry, to determine the proportions of A) total MAIT cells within the CD3^+^ T cell compartment, B) CD8^+^ T cells within the CD3^+^ MAITs and C) CD4^+^ T cells within the CD3^+^ MAIT compartment. PBMC samples were analysed by four separate flow cytometry staining panels: in common, PBMCs were stained extracellularly for immune cell profiles, then specific stain methods were engaged. “AE”: Activation/Exhaustion markers; “Func”: functional markers; “TF”: transcription factor markers; “FUNCin_gm”: functional marker staining following incubation overnight in culture medium. Each dot represents the average value across all the samples collected at different time points for one individual study participant. Mean values and SD were plotted in graphs. Datasets were compared using the Mann-Whitney test for non-parametric data. Statistical significance was set at p < 0.05.

### T cell and MAIT memory subsets show signs of early senescence in ME-MM

We next investigated whether there were differences in the memory and differentiation states of T cells and MAITs from people within the ME-MM and ME-SA groups, by measuring naïve and memory cell markers (CD45RA, CCR7) and differentiation markers (CD28, CD57). Naïve cells were defined as CCR7^+^CD45RA^+^, CCR7^+^CD45RA^-^ as central memory (CM), CCR7^-^CD45RA^-^ as effector memory (EM) and CCR7^-^ CD45RA^+^ as terminally re-expressing CD45RA effector cells (TEMRA). There were no significant differences in frequencies of naïve/memory subsets in CD4^+^ or CD8^+^ T cells between the ME-MM and ME-SA groups, nor were there any differences in the frequencies of CD57^-^CD28^+^, CD57^-^CD28^-^, CD57^+^CD28^-^ or CD57^+^CD28^+^ subsets (Supplemental Figure 4). Next, we analysed each CD4^+^ and CD8^+^T cell naïve/memory subset by CD57 and CD28 expression. In CD4^+^ T cells, >95% of the naïve cells were CD57^-^CD28^+^, and the frequency of this phenotype reduced through the Naïve, CM, EM then TEMRA cell compartments, whereas the frequency of the CD57^+^CD28^-^ subset, which associated with senescence (33–35), gradually increased from the Naïve cells through CM then EM to TEMRA, although there were no significant differences between two clinical groups (Supplemental Figure 5). However, the frequency of the CD57^-^CD28^-^ subset, which is considered a marker of early senescence alongside highly differentiated phenotype (34), was significantly higher in the CD4^+^ CM T cell subset in the ME-MM group compared to the ME-SA group, with a non-significant trend (p=0.0899) towards higher frequency also in the CD4^+^ EM T cells in the ME-MM group. Moreover, a similar pattern was also seen in CD8^+^ T cells, with the frequency of CD57^-^CD28^-^ cells in the CD8^+^ EM and CD8^+^ TEMRA T cell subsets being significantly elevated in ME-MM compared to ME-SA (p=0.012 in CD8^+^ EM, p=0.019 in CD8^+^ TEMRA) (Figure 2B). CD3^+^ MAIT cells reportedly display an effector memory cell phenotype (36, 37). Here, we found MAIT cells expressed CD28, but not CD57 on their surface. Interestingly, the frequency of CD28^+^ MAIT cells was significantly reduced in ME-MM compared to the ME-SA group (p=0.017) (Figure 2C). The majority of CD8^+^ MAITs expressed CD28, without no significant difference between the two clinical groups; however, CD4^-^CD8^-^ MAIT cells had reduced frequency of CD28 expression in the ME-MM group compared to those from people with ME-SA (p=0.007) (Figure 2C).

**Figure 2.**
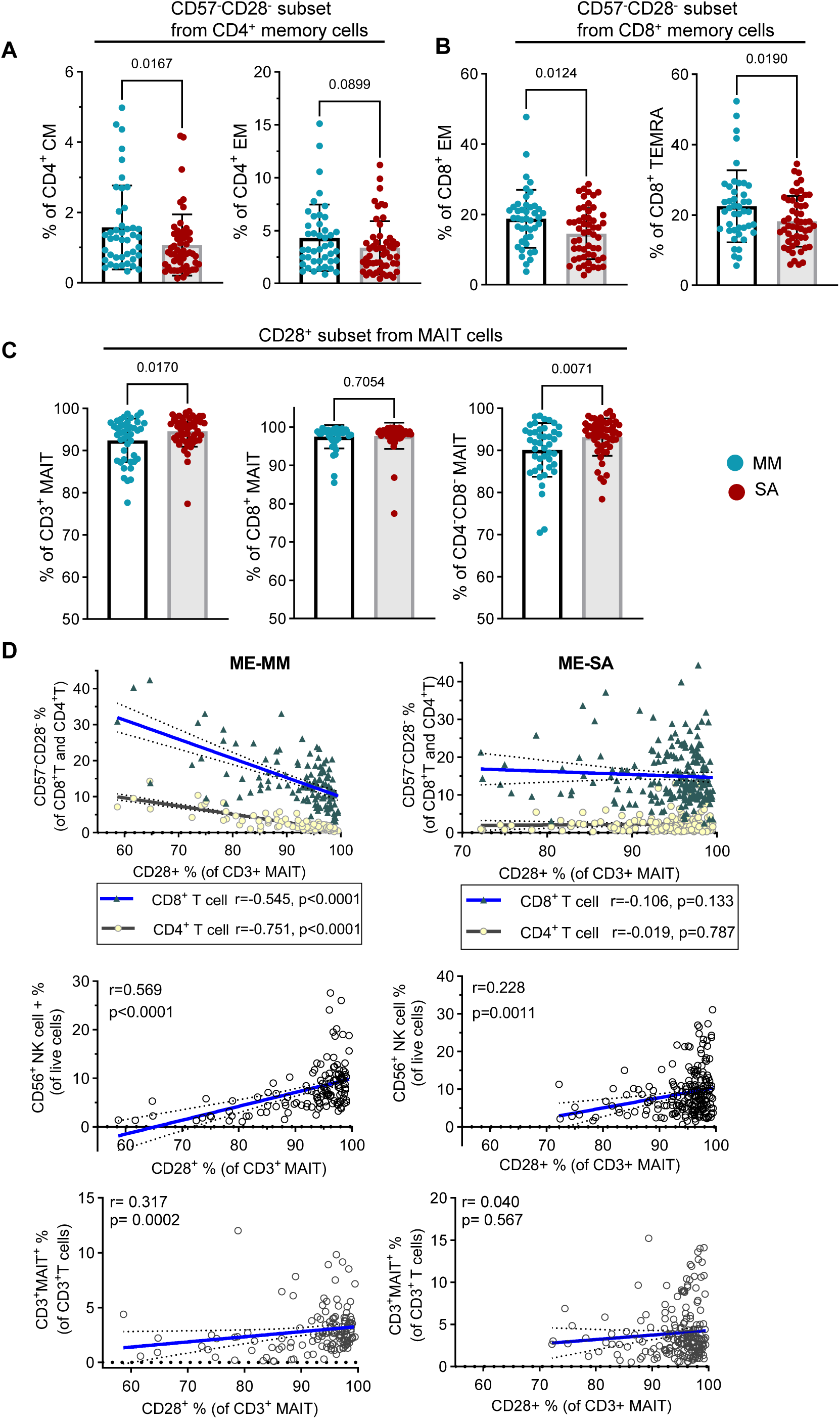
Differential expression of CD57^-^CD28^-^ subsets in memory T cells and CD28^+^ MAIT cells in mild/moderate (n=43) and severe ME/CFS (n=53). PBMC samples were analysed using the ‘memory/differentiation panel’ comprising CD45RA, CCR7, CD57, and CD28. A) CD57^-^CD28^-^subsets (%) in central memory (CM) CD4^+^T cells (left) and effector memory (EM) CD4^+^T cells (right). B) CD57^-^CD28^-^ subsets (%) in EM CD8^+^T cells (left) and terminally differentiated (TEMRA) CD8^+^T cells (right). C) CD28+ subset (%) in total CD3^+^MAIT cells (left), in CD8^+^ MAIT cells (middle) and CD4^-^CD8^-^ MAIT cells (right). Each dot represents the average value across all the samples collected at different time points for one individual. Mean values and standard deviation (SD) are shown. D) Correlation analysis of frequencies of CD28^+^ MAIT cells and CD57^-^CD28^-^ subsets in CD4^+^T cells and CD8^+^T cells (upper), NK cells (middle), and CD3^+^ MAIT cells (bottom). A total 335 samples (n=132 for MM, n=203 for SA) were used for the Spearman correlation test; the dashed line represents the 95% confident interval.

We then performed correlation analyses which showed differences between the ME-MM and ME-SA groups in the association between the frequency of CD28^+^ MAIT cells and of T cell CD57^-^CD28^-^ subsets: there was a negative correlation in the ME-MM group for both CD4^+^ (r= −0.751, p<0.0001) and CD8^+^ (r=-0.545, p<0.0001) CD57^-^CD28^-^ T cells and CD28^+^ MAITs (Figure 2D). These observations were not observed in people with ME-SA. As MAITs cells act as innate-like T cells in response to microbial infection, we performed a correlation analysis between the frequency of NK cells, another innate immune cell type, and MAITs to determine whether these innate immune cells’ appearance together would affect the first line of defence in microbial infection, and found that the frequency of CD28^+^ MAITs was moderately correlated with the frequency of NK cells (r=0.569, p<0.0001) in ME-MM. Overall, this may indicate that reduced frequency of CD28 expression in MAITs from ME-MM is associated with higher frequency of CD57^-^CD28^-^ T cell subsets and lower frequency of NK cells.

### Granzyme and/or perforin expressing CD8^+^ T cells and MAITs are reduced in people with ME-SA, ex vivo and following stimulation

To determine the functional capacity of lymphocytes in people with ME-MM or ME-SA, we measured intracellular cytotoxic molecules (granzyme B and perforin) and cytokines (IFN-γ and IL-17) in T cells and NK cells. Intracellular cytokines were not detectable in *ex vivo* T cells (data not shown). Most NK cells contained granzyme B and perforin (granzyme B: mean 75.4% in ME-MM, 73.9% in ME-SA; perforin: 75.3% in ME-MM, 74.4% in ME-SA) (Supplemental Figure 6A). Although there was no difference in the frequency of cytotoxic NK cells between the ME-MM and ME-SA groups, there was a reduced amount of granzyme B in NK cells in the ME-SA group, measured by MFI, although not of perforin (p=0.025 in granzyme B expression, p=0.340 in perforin) (Figure 3A). There were no differences in the frequencies of granzyme B and perforin expressing cytotoxic CD8^+^ T cells and CD4^+^ T cells between the ME-MM and ME-SA groups (Supplemental Figure 6B and 6C). However, the MFI of both granzyme B and perforin in CD8^+^ T cells was significantly reduced in the ME-SA group (p=0.021 in granzyme B expression, p=0.023 in perforin) (Figure 3B). Granzyme B was detected at low frequencies in MAITs in *ex vivo* PBMC samples, although significantly more highly in the ME-SA than ME-MM group (mean 0.46 % in ME-MM, mean 1.25% in ME-SA) (Figure 3C). However, the granzyme B-positive frequency was significantly increased after incubation for 18 hr in growth medium alone in ME-MM, but not in ME-SA (ME-MM; 2.10%, ME-SA; 1.46 %, p=0.002) (Figure 3C; Supplemental Figure 7). After *in vitro* culture with the stimulation cocktail comprised of PMA and ionomycin, the frequencies of intracellular granzyme B-expressing MAITs were significantly increased in both groups (p<0.0001 for both), with a significantly greater up-regulation in the ME-MM group (mean 14.94 %) compared to the ME-SA group (11.73 %) (p=0.047) (Figure 3C). A moderate proportion of MAITs expressed perforin *ex vivo*: the frequency of perforin-expressing MAITs was significantly lower in the ME-SA group (17.80 % in ME-MM, 12.72 % in ME-SA) (p=0.001) (Figure 3D). Furthermore, we investigated the main source of MAIT subsets which expressed perforin, and identified the CD4^-^CD8^-^ double negative MAIT subset as the major contributor, rather than the expected CD8^+^ MAIT subset, in both clinical groups (p<0.001 in MM, p=0.038 in SA) (Figure 3D). Together, these data show that CD8^+^ T cells and MAITs from people with ME-SA had a reduced capacity to express the cytotoxic mediators perforin and granzyme.

**Figure 3:**
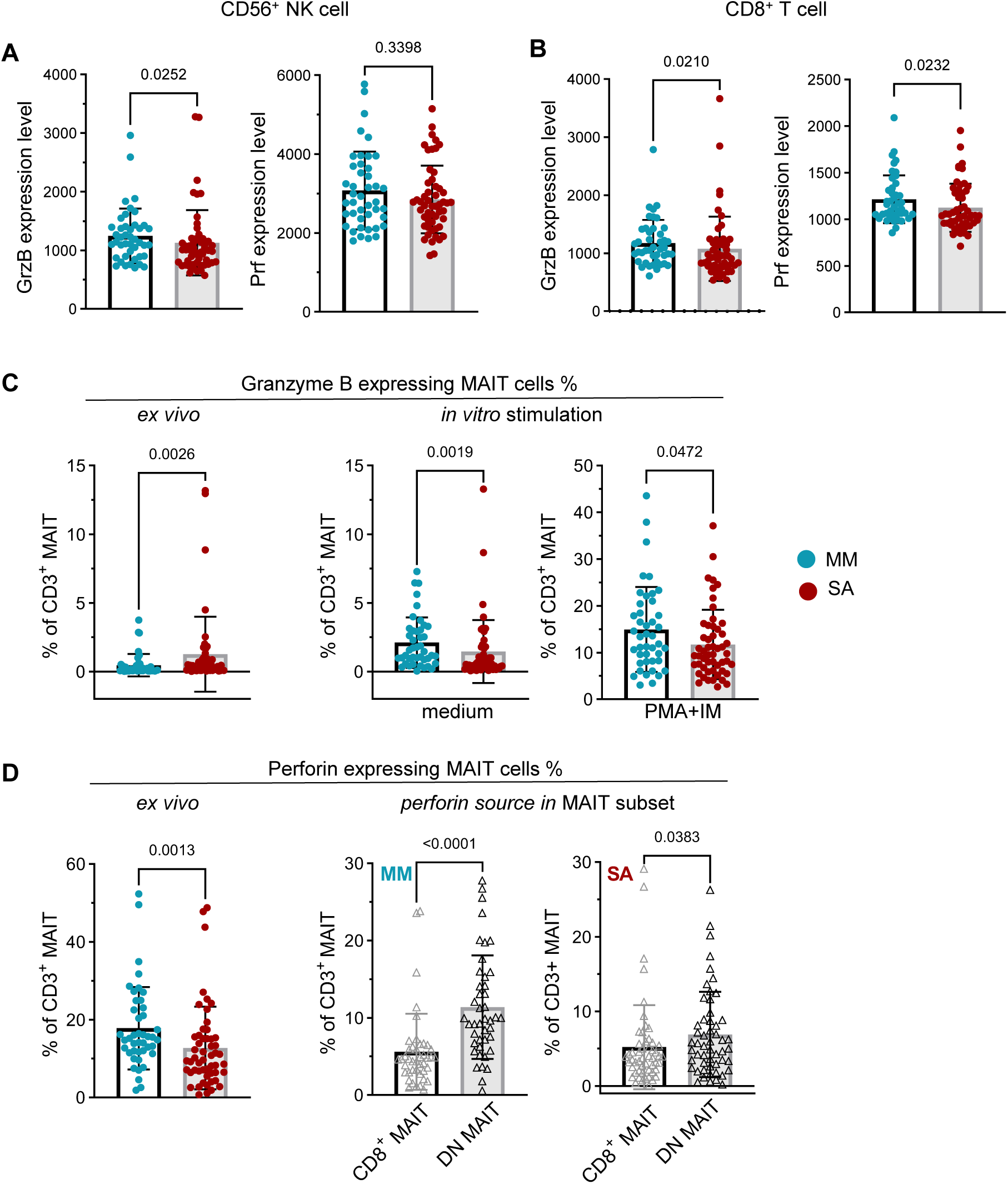
*Ex vivo* and *in vitro* analysis of expression of cytotoxic mediators in cytotoxic cells (NK cell, CD8^+^T cells and MAITs) from people with mild/moderate (n=43) and severe ME/CFS (n=53). Thawed PBMC samples were stained for intracellular Granzyme B (GrzB) and perforin (Prf). Median fluorescence intensity (MFI) of GrzB (left) and Prf (right) was measured in the cytotoxic molecule-expressing NK cell population (A) and CD8^+^T cell population (B). C) The frequencies of GrzB-expressing CD3^+^ MAITs were compared between ME-MM and ME-SA in PBMC directly after thawing (left), resting overnight in culture medium (middle) and following 5 hours stimulation with PMA and ionomycin after incubation overnight (right). D) Prf expression in CD3^+^MAITs was compared between ME-MM and ME-SA groups in thawed PBMC (left), and the main source of the Prf expression in CD3+MAIT cells was analysed in CD8^+^ MAITs cells and CD4^-^CD8^-^ DN MAITs in the ME-MM (middle) and ME-SA (right) groups. Each dot represents the average value across all the samples collected at different time points for one individual. Mean values and SD are shown. Datasets were compared using the Mann-Whitney test for non-parametric data or the t-test for parametric data, with p<0.05 deemed significant.

### People with ME-MM exhibited stronger correlation between cytotoxic marker expression and senescent T cells and DN MAITs

To explore the potential interaction between the frequencies of cytotoxic T cell subsets and the various T cell differentiation subsets, especially the senescent CD57^+^CD28^-^ subsets, we next performed correlation analyses between the frequencies of different cell populations on all the samples (n=335) or by the two clinical groups separately (n=132 for ME-MM, n=203 for ME-SA). Using the data acquired using the ‘memory/differentiation’ and ‘function’ flow cytometry staining panels, we found that the frequency of cytotoxic marker-positive CD8^+^ T cells was strongly correlated with the frequency of CD57^+^CD28^-^ CD8^+^ T cells (r=0.939 for GrzB, r=0.684 for Prf, Supplemental Figure 8A). The correlation strengths were similar in the two clinical groups. This pattern was also observed in CD4^+^ T cells, although cytotoxic effector molecules were less frequent in CD4^+^ T cells, as expected (Supplemental Figure 8B). The correlation analysis showed that samples with a lower CD28^+^ frequency in MAITs cells had a higher perforin-positive MAIT frequency (r= − 0.209, p=0.0001; Supplemental Figure 8C); furthermore, the extent of correlation was much stronger in the ME-MM group compared to the ME-SA group (r=-0.429, p<0.0001 for MM, r=-0.044, p=0.529 for SA; Supplemental Figure 8C). There was an even stronger association in the DN MAIT subset: the frequency of DN MAITs was positively correlated with the frequency of perforin^+^ MAIT cells across all the samples (r=0.393, p<0.0001 for the total sample set), while the ME-MM clinical group had higher correlation coefficients than the ME-SA (r=0.612, p<0.0001 for ME-MM; r=0.234, p=0.0008 for ME-SA; Supplemental Figure 8D). In agreement with these results, there was a negative correlation between the frequencies of CD28^+^ MAITs and DN MAITs (Supplemental Figure 8E). These findings may suggest that the samples with a higher frequency of DN MAIT cells have more cytotoxic capacity, with a more frequent appearance of the CD28^-^ population in MAITs: this is in accordance with our finding that the ME-MM group showed a non-significant trend towards higher frequency of DN MAITs (data not shown) as well as increased cytotoxic marker expression in MAITs and downregulation of the CD28^+^ MAIT subset.

### T cells and NK cells from people with severe ME/CFS exhibit enhanced activation status

Next, we investigated the expression of the activation markers CD69 and CD38, as well as the exhaustion marker, PD-1, in *ex vivo* PBMCs. First, we examined the association between CD69 expression on different cell types. We found a moderate and highly significant (p<0.0001) correlation between the frequency of MAITs expressing CD69 and the frequency of NK cells, CD4^+^ T cells and CD8^+^ T cells expressing CD69 across all the samples (Spearman r=0.697 for CD69^+^NK cells, r=0.508 for CD69^+^CD4^+^ T cells, r=0.649 for CD69^+^CD8^+^ T cells, Figure 4A). Comparing the two groups, we found people with ME-SA had an elevated frequency of CD69 expression on T cell subsets, including CD4^+^ and CD8^+^ T cells and MAITs cells, compared to people with ME-MM (p=0.026 in CD4^+^ T cell, p=0.023 in CD8^+^ T cells, p=0.019 in MAIT cells) (Figure 4B). Moreover, the frequency of CD38^+^CD69^+^ cells within the CD4^+^ and CD8^+^ T cells and in MAITs was also significantly increased in ME-SA (p=0.007 in CD4^+^ T cell, p=0.012 in CD8^+^ T cells, p=0.001 in MAIT cells) (Figure 4C). Together, these data reveal a global increased activation status of lymphocytes in severe ME/CFS compared to mild/moderate illness. We also found the PD-1 expression level (MFI) from the PD-1^+^ subsets of CD4^+^ T cells, CD8^+^ T cells, MAITs and NK cells was lower in ME-SA (p=0.020 in CD4^+^ T cells, p=0.030 in CD8^+^ T cells, p=0.0004 in MAITs, p=0.0004 in NK cells) (Figure 4D), although there was no significant difference in frequency of PD-1 expressing T cells and NK cells (data not shown). Furthermore, we also analysed another activation indicator, the co-expression of human leukocyte antigen (HLA)-DR and CD38 in CD4^+^ T cells and CD8^+^ T cells, which are considered to be expressed during viral infection (38–40). As shown in Figure 4E, there was a significantly higher frequency of co-expression of HLA-DR and CD38 on CD4^+^ and CD8^+^ T cells in ME-MM compared to the ME-SA group (p=0.018 in CD4^+^ T cell, p=0.021 in CD8^+^ T cells).

**Figure 4.**
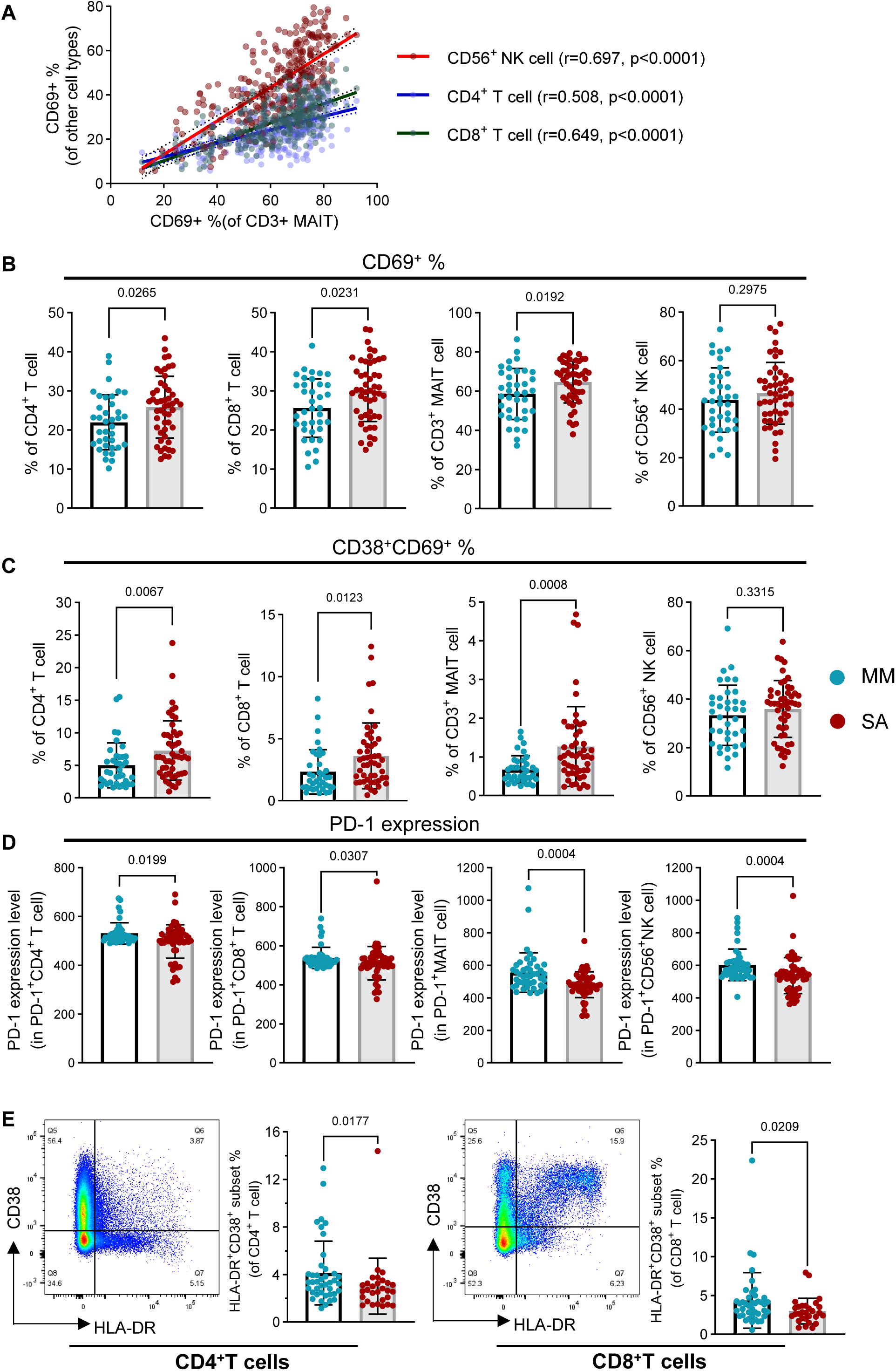
Frequencies of activation/exhaustion marker expression in T cells and NK cells from people with mild/moderate (n=43) and severe ME/CFS (n=53). *E vivo* PBMC were stained for surface CD69, CD38 (both as activation markers) and PD-1 (as an exhaustion marker). A) Correlation analysis was performed between CD69^+^ MAITs cells and other cell types (CD69^+^ NK cells, CD69^+^CD4^+^T cells, CD69^+^CD8^+^T cells). A total of 335 samples (n=132 for ME-MM, n=203 for ME-SA) were analysed using the Spearman correlation test. B) The frequencies of CD69^+^ T cell subsets (CD4^+^ T cells, CD8^+^ T cells and MAITs) and NK cells were compared between ME-MM and ME-SA groups. C) Comparison of CD38^+^CD69^+^ frequency in T cells (CD4^+^ T cells, CD8^+^ T cells and MAITs) and NK cells between ME-MM and ME-SA groups. D) Comparison of PD-1 expression (median fluorescence intensity) in PD-1^+^ subsets in T cells (CD4^+^ T cells, CD8^+^ T cells and MAITs) and NK cells. E) Representative pseudocolour plots of HLA-DR^+^CD38^+^ staining, and comparison of the frequency of HLA-DR^+^CD38^+^ T cell subsets between ME-MM (n=41) and ME-SA (n=29) in CD4^+^ T cells (left) and CD8^+^T cells (right). Each dot represents the average value across all the samples collected at different time points for one individual. Mean values and SD are shown, with p<0.05 deemed significant.

### Proinflammatory cytokine production after stimulation is elevated in ME-SA

We then examined the frequency of intracellular cytokine expression (TNF, IFNγ and IL-17), along with CD69, in T cells following stimulation with PMA and ionomycin for 5 hours. As shown in Figure 5A, most T cell subsets expressed CD69 after *in vitro* stimulation. The frequencies of CD69^+^CD4^+^ T cells and CD69^+^ MAITs were significantly higher in ME-SA (p=0.046 in CD4^+^ T cell, p=0.002 in MAITs) (Figure 5A), consistent with the *ex vivo* PBMC result. Overall, IL-17 was rarely detected (Figure 5B), however TNF was produced by CD4^+^ T cells (48.7% in ME-MM, 53.8% in ME-SA, p=0.061), CD8^+^ T cells (30.0% in ME-MM, 35.5% in ME-SA, p=0.080) and MAITs (66.4 % in ME-MM, 79.0 % in ME-SA, p=0.002) with higher frequencies in ME-SA (Figure 5C). The frequencies of IFNγ-producing CD8^+^ T cells and MAITs were elevated in people with ME-SA compared to ME-MM (p=0.022 in CD8^+^ T cells, p=0.002 in MAIT cells) (Figure 5D). Moreover, CD8^+^ T cells from people with ME-SA had elevated frequencies of TNF, IFNγ and IL-17 production (p=0.080 in TNF, p=0.022 in IFNγ, p=0.012 in IL-17), and MAITs from people with ME-SA had significantly elevated frequencies of TNF (p=0.0002) and IFNγ (p=0.002) production. Furthermore, IFNγ and TNF co-producing bifunctional MAIT cell frequency was significantly elevated in the ME-SA group (56.1%) compared to the ME-MM group (48.4 %) (p=0.048) (Figure 5E), whereas the proportion of IFNγ^-^ TNF^-^ double negative MAITs was reduced (p=0.001). We also found that MAIT cell activation by PMA and ionomycin changed the proportion of subsets compared to the non-stimulated condition, with an increase of DN MAITs (from 33.1% to 43.9 % in ME-MM and from 23.9% to 35.1 % in ME-SA) and a reduction of CD8^+^ MAITs (from 63.8 % to 46.5 % in ME-MM and from 74.3 % to 58.9 % in ME-SA) in both groups (Supplemental Figure 9).

**Figure 5:**
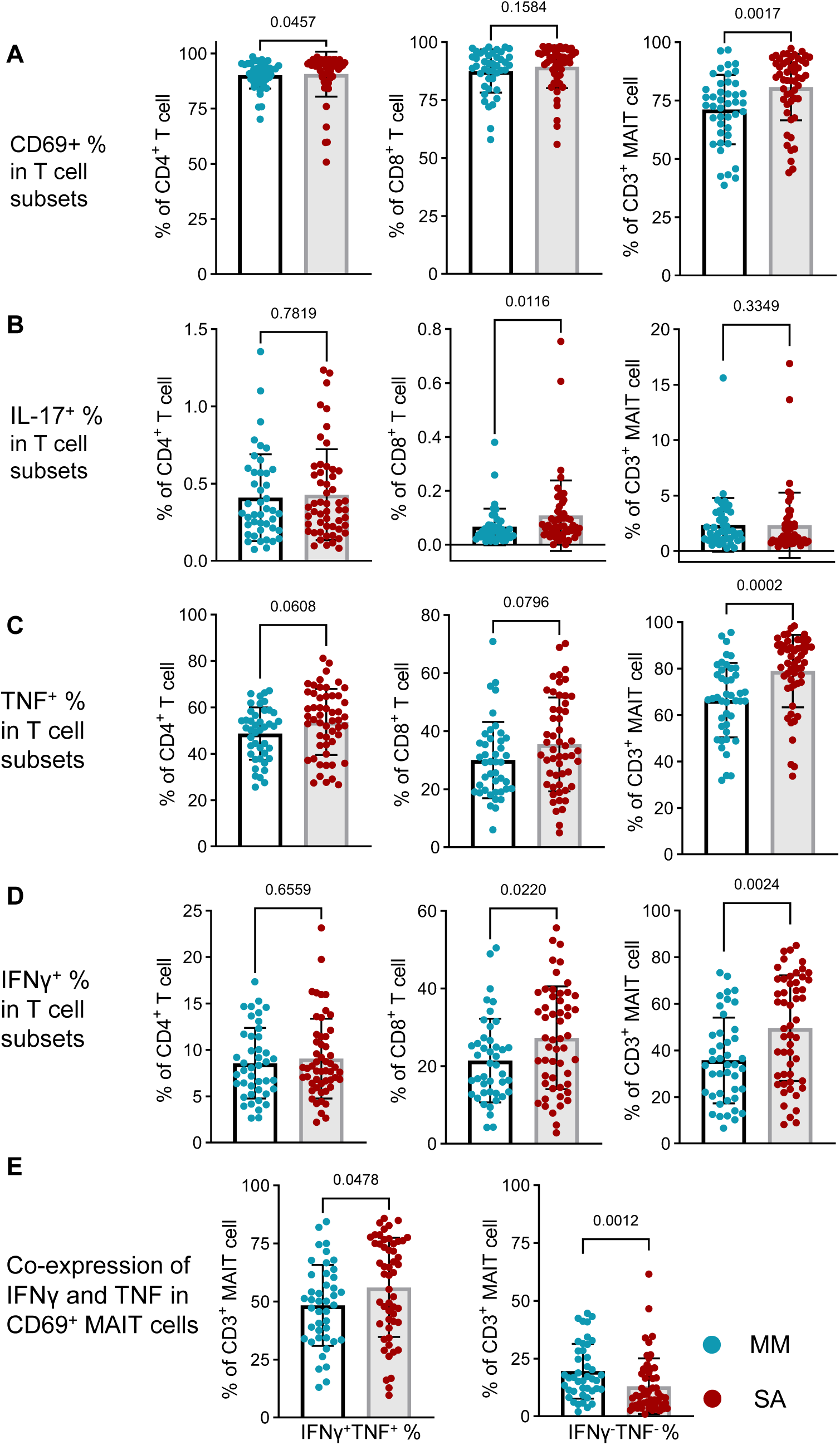
*In vitro* stimulation of PBMC and functional analysis of T cells from people with mild/moderate (n=43) and severe ME/CFS (n=53). Thawed PBMCs were incubated overnight, and then stimulated with PMA and ionomycin for 5 hours. Then the stimulated cells were stained for surface T cell markers (CD4, CD8, MAIT) along with the activation marker CD69, then intracellular cytokines (TNF, IFNγ and IL-17). The unstimulated cells were stained in parallel. A) The CD69 expression was compared between two groups in three T cell subsets. B-D) The frequencies of CD4^+^ T cells, CD8^+^ T cells and MAITs expressing IL-17 (B), TNF (C) and IFNγ (D) were compared between the ME-MM and ME-SA groups. E) TNA and IFNγ co-expressing CD69^+^ MAITs (left) and TNA^-^ and IFNγ^-^ non-expressing CD69^+^ MAITs were compared between the clinical groups. Each dot represents the average value across all the samples collected at different time points for one individual. Mean values and SD are shown. Datasets were compared using the Mann-Whitney test for non-parametric data or the t-test for parametric data, with p<0.05 deemed significant.

To investigate the intranuclear mechanism driving higher cytokine production in the ME-SA group, we examined the expression of transcription factors eomesodermin (EOMES) and T-box transcription factor (T-bet) in T cell subsets and NK cells in *ex vivo* PBMCs following intranuclear staining. EOMES and T-bet are known to drive a type 1 response, and T-bet is an essential transcription factor for optimal IFNγ production as well as NK cell maturation (41–43). We observed the ME-SA group had a non-significant trend towards higher frequency of EOMES-expressing CD8^+^T cells and no difference in the frequency of T-bet expressing CD8^+^ T cells compared to the ME-MM group. Both the frequency and MFI of T-bet expression in MAITs were significantly increased in the ME-SA (p=0.003 in frequency of T-bet, p=0.001 in MFI of T-bet) whereas there was no difference in EOMES expression (Figure 6). These results indicate that elevated T-bet expression in MAITs cells and more frequent EOMES expressing CD8^+^ T cells in circulating PBMC from ME-SA may play a boosting role to produce more proinflammatory cytokines after *in vitro* stimulation. In addition, T-bet expression level (MFI) in T-bet^+^ NK cells was significantly elevated in ME-SA (p=0.044), although there was no significant difference in the frequency of T-bet^+^ NK cells because T-bet was commonly detectable in NK cells in both groups (Supplemental Figure 10).

**Figure 6:**
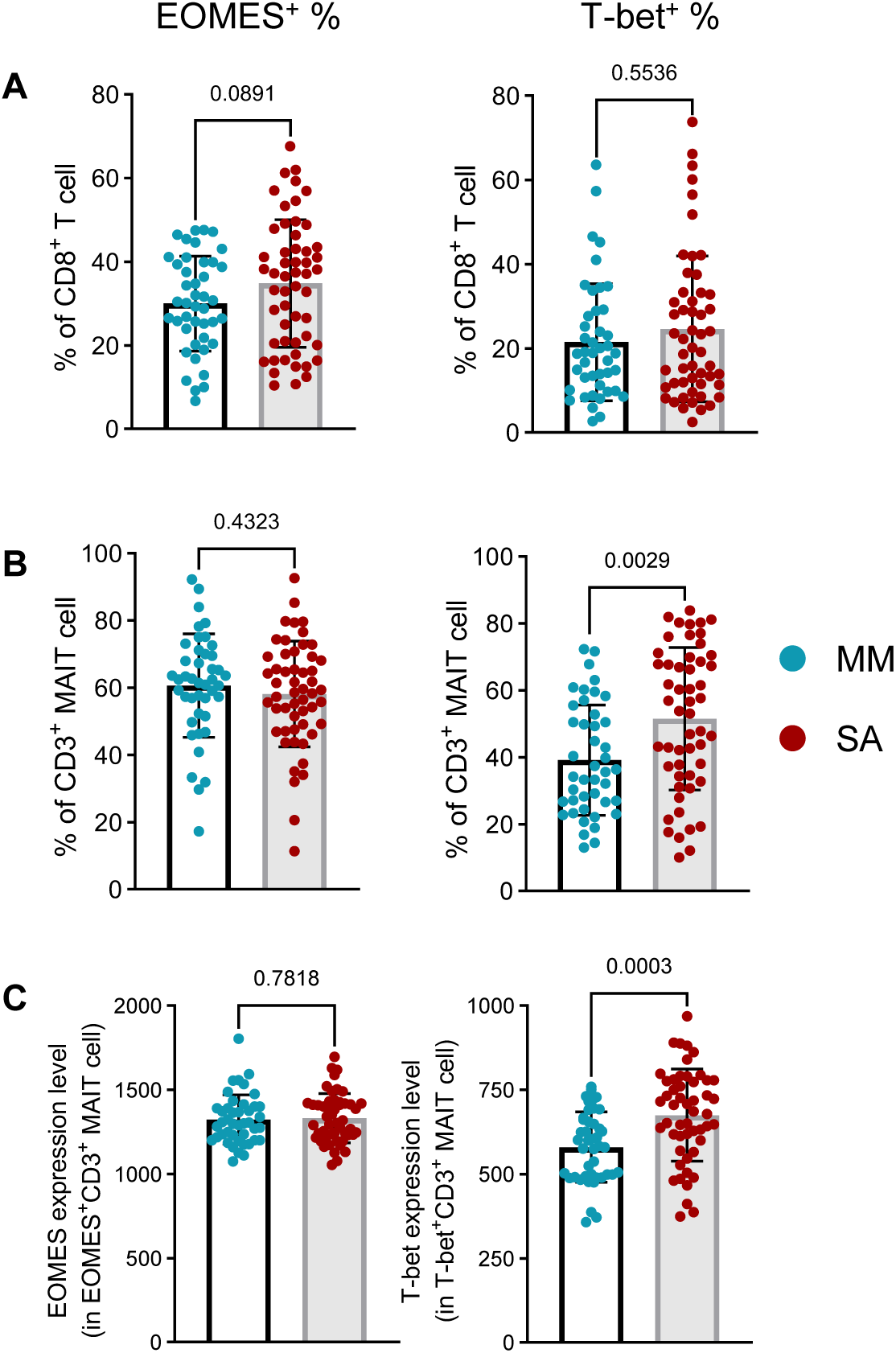
Eomes and T-bet expression in CD8^+^T and MAIT cells from people with mild/moderate (n=43) and severe ME/CFS (n=53). The frequency of expression of the EOMES and T-bet transcription factors was quantified by intranuclear staining after the cell phenotype extracellular staining, for CD8^+^ T cells (A), along with the frequency and MFI expression in MAITs (B, C). Each dot represents the average value across all the samples collected at different time points for one individual. Mean values and SD are shown. Datasets were compared using the Mann-Whitney test for non-parametric data or the t-test for parametric data, with p<0.05 deemed significant.

### T cell activation, cytotoxicity and senescence phenotypes can discriminate mild/moderate and severe ME/CFS

As there were significant differences observed in the expression of activation markers, proinflammatory cytokines, cytotoxicity effector molecules and immunosenescence markers between people with ME-MM and ME-SA, we next sought to determine whether combinations of these markers could be used to predict the disease severity classification. The majority of the ME-MM participants were separated from the majority of the ME-SA participants in principal component analyses based on combinations of cell markers in CD8^+^ T cells or in MAITs separately, with enhanced separation evident when the expression profiles of both CD8^+^ T cells and MAITs were combined (Supplemental Figure 11). These cell markers had moderate capacity to discriminate between the ME-MM and ME-SA clinical groups in Receiver Operating Characteristic curve analyses, with an Area Under the Curve of 0.7141 (p=0.0007) for the CD8^+^T cell markers and 0.758 (p<0.0001) for the MAITs. The combination of the CD8^+^ and MAIT markers had an AUC of 0.7843 (p<0.0001) (Supplemental Figure 11F), indicating that the activation, inflammation, cytotoxicity and immunosenescence markers have the capability to discriminate between people with mild/moderate or severe ME/CFS disease.

## Discussion

In this study we have demonstrated clear differences exist in immune cells in people with mild/moderate compared with severe ME/CFS. These differences imply there are different processes pathophysiological processes occurring in the two clinical groups. We focused on cytotoxic cell phenotype and function, including NK cells, CD8^+^ T cells and MAITs, as they have all previously been implicated in ME/CFS (12, 15, 44–47). We found that people with mild to moderate ME/CFS symptoms had more evidence of early-senescence in their memory T cell subsets and MAITs, shown by higher frequencies of CD57^-^CD28^-^ subsets within central memory CD4^+^ T cells, effector memory and TEMRA CD8^+^ T cells and lower frequencies of the CD28^+^ subset in MAITs. Cells from ME-MM participants also showed increased granzyme B and/or perforin expression in cytotoxic cells from *ex vivo* and *in vitro* stimulated PBMCs. Conversely, the frequencies of activated CD69^+^ T cells, as well as CD38^+^CD69^+^ T cells, were increased in people severely affected with ME/CFS, and CD69 was more highly enhanced following stimulation in the ME-SA group. Pro-inflammatory cytokines IFNγ, TNF and IL-17 were more highly expressed in stimulated cells from severely affected participants. As we have described previously (18), this group also had elevated frequencies of circulating MAITs, more of which were CD8^+^ MAITs, whereas CD4^+^ MAITs cells were significantly lower in frequency than in the ME-MM group. Importantly, strong correlations were observed across different cells and cell subsets, for cytotoxic effector molecules, activation markers and immunosenescence markers, indicating a systemic T/NK cell response rather than a specific unique lymphocyte subset being affected in ME/CFS. These differences in CD8^+^ T cell and MAIT markers had the potential to distinguish between the clinical groups.

It is still not well established if mild/moderate and severe cases of ME/CFS represent a spectrum of clinical phenotypical expressions of the same pathophysiological processes, or if they rather largely relate to distinct processes. With a paucity of studies investigating more severely affected people living with ME/CFS, our study adds important information on immunological differences between these groups. Treatment approaches may therefore need to vary across groups. Our results also suggest that biomarkers can be developed which classify to which sub-group a person with ME/CFS belongs, to be used prognostically and to aid treatment choices when new treatments become available.

MAIT cells were upregulated in frequency in ME-SA compared to ME-MM study participants. MAITs are an innate-like T cell subset which play an antimicrobial role by recognising microbe riboflavin derivates presented in an MHC-related protein 1-dependent manner, to produce proinflammatory cytokines as well as cytotoxic molecules. They are found in human peripheral blood at varying frequency (1-10% of T cells) as well as in tissues such as the liver and the intestines (48, 49), and are generally significantly reduced or depleted in blood in in bacterial and viral infectious diseases, metabolic disorders, and chronic inflammatory and autoimmune diseases (50). It is not known why they are increased peripherally in severe ME/CFS: of note, MAIT proliferation has been shown to occur in response to *E.coli* (51, 52) and in people latently-infected with *Mycobacterium tuberculosis* (53), and it is plausible that MAIT proliferative capacity is enhanced in ME-SA. Alternatively, there could be dysregulation of tissue-homing chemokine receptor and integrin expression in ME-SA: a recent report demonstrated that normally MAITs are recruited to acutely inflamed intestine via the differential expression of CXCR6 and α4β7 (54), and disruption of this trafficking in ME-SA could be another explanation for increased MAITs seen in ME-SA in this study. Furthermore, the gut microbiome composition which is affected by ME/CFS (55), affects the abundance of MAITs (56), an association that warrants further investigation.

MAIT subsets, determined by CD4 and CD8 coreceptor expression, are known to be functionally distinct. CD8^+^ MAITs are reportedly more cytotoxic, with a stronger type 1 response phenotype and different transcription profile, in comparison with the developmentally related CD4^-^CD8^-^MAIT (DN MAIT) cells (57) and with CD4^+^ MAITs (58), although CD4^+^ MAITs cells express increased CD25 and IL-2 signalling (58). In our study, we found the frequencies of MAIT subsets, as proportions of total MAITs (defined as CD3^+^CD161^++^TCRVα7.2^+^MR1-5-OP-RU^+^), were different between the ME-MM and ME-SA clinical groups. In both groups, CD8^+^ MAITs were most dominant, followed by the DN MAITs, CD4^+^ MAITs and then CD4^+^CD8^+^ DP MAITs, but the frequency of CD8^+^ MAITs was increased in ME-SA compared to ME-MM, whereas DN MAITs and CD4^+^ MAITs cell were higher in ME-MM. As CD4^+^ MAITs cells were detected at a low frequency of total MAITs (< 5 % of CD3^+^MAIT cells), we focused on the CD8^+^ MAIT cells and DN MAIT cells for further analysis. We also found that MAIT cell activation by PMA and ionomycin induced an increase of DN MAITs and a reduction of CD8^+^ MAITs, possibly due to a loss of CD8 on the surface of CD8^+^ MAITs, as well as significantly higher perforin expression in DN MAITs *ex vivo* in both clinical groups. Together, the MAIT cell data indicate a reduction in cytotoxic function in MAITs in severely affected individuals, associated with an increase in CD8^+^ MAITs and decrease in DN MAITs, in contrast to existing non-ME/CFS based literature (57).

We identified an expansion in the proportions of CD4^+^ and CD8^+^ memory T cells and MAITs which exhibited early stages of immunosenescence, based on the loss of CD28 expression but absence of CD57 expression, in the mild/moderate ME/CFS group. CD28 is a necessary costimulatory molecule for T cell activation, and the CD28^-^ T cell population is regarded as having a cytotoxic or regulatory phenotype (59). Using microarray analysis, Pangrazzi *et al* reported that CD28^-^CD57^-^ CD8^+^ T cells showed early senescence characteristics with reduced production of IFNγ and TNF production (60). The CD28^-^ subset in CD8^+^T cells is expanded by chronic viral infection such as human cytomegalovirus, Epstein-Barr virus and human parvovirus B19, which have been implicated in ME/CFS pathogenesis (35, 61, 62). We also observed significantly diminished CD8 staining intensity (reduced MFI) on CD8^+^ T cells in ME-MM compared with ME-SA. Reduced CD8 expression level occurs transitorily in bacterial and viral infection (63), and is accompanied by enhanced cytotoxic effector function (64). Along with the CD8 down-regulation, we observed an increase in granzyme B and perforin expression, determined from MFI data, in CD8^+^ T cells and NK cells, in ME-MM. Taken together, these findings may suggest that the appearance of senescent and highly differentiated subsets in T cells and increased cytotoxicity as well as the lower CD8 MFI is caused by frequent exposure to antigens. In this regard, we have recently reported that human herpesviruses 6B and 7 DNA in saliva correlates with symptom severity in ME/CFS (65), and it is plausible that herpes virus reactivation stimulates T cells, driving them to cytotoxicity and senescence.

In contrast, the early leukocyte activation marker CD69 was more highly expressed in the ME-SA group, on *ex vivo* CD4^+^ and CD8^+^ T cells and on MAITs, and was also more upregulated by stimulation with PMA and ionomycin in people with ME-SA than ME-MM. Thus T cells from people severely affected with ME are more readily activated both in the circulation and *in vitro*. Upregulated CD69-expressing immune cells in blood have been reported in autoimmune disorders, such as Graves’ disease (45) and psoriasis (66, 67), although CD69-expressing cells have been suggested to either promote (68) or reduce disease progression (69) in models of systemic lupus erythematosus. We also observed that co-expression of another activation marker CD38 with CD69 was increased on CD4^+^ and CD8^+^ T cells and MAITs in ME-SA: a higher frequency of CD38^+^CD69^+^T cells has also been reported in acute hepatitis E virus infected patients, compared to those from resolving-phase patients (70). The pathophysiological mechanism underlying the enhanced frequency of circulating CD69^+^CD38^+^ T cells in ME-SA warrants further investigation. Notably, increased frequencies of IFNγ-expressing cells, either alone or in combination with TNF production, were found in T cell subsets following *in vitro* stimulation in the ME-SA group, associated with an increased T bet expression in MAITs. The enhanced frequency of IFNγ^+^TNF^+^CD69^+^ CD8^+^ MAITs we discovered indicates an ongoing pro-inflammatory response in ME-SA. Activated CD8^+^ MAITs are able to respond rapidly as polyfunctional effectors either TCR dependently or independently (71).

The strengths of our study include our focus on a very well characterised cohort. All participants completed a Symptoms Assessment form to confirm case definition compliance and study eligibility before they were accepted onto the study. Clinical assessment and additional validated questionnaires allowed further characterisation of cases by clinical phenotype and disease severity. Clinical assessment data were collected using standard equipment by an experienced research nurse trained in the study’s clinical assessment protocol. We were also able to include severely affected individuals in our study recruited using home visits: this group is often excluded from research studies. The participants provided blood samples at repeated time points, enabling us to find consistent trends between the clinical sub-groups. The study was designed to determine the immunological differences between people who are mild-to-moderately and those severely affected with ME/CFS, and a limitation of the study was that no healthy controls were included in the follow-up study. We have however previously reported differences in MAIT frequencies and CD8^+^ T cell subsets between healthy donors and PWME (18), and this current study greatly expands this by comparing averaged values from individuals through different timepoints between the two clinical groups, and by performing a detailed characterisation of cell differentiation, senescence and effector cell function. Further prospective longitudinal analyses with healthy controls will be required to facilitate the development of new treatments and diagnostic signatures, and future larger scale studies should include training, test and validation cohorts for biomarker verification.

In conclusion, our results elucidate that people with ME/CFS with different symptom severity had different immune profiles in T cells in aspects of activation, differentiation and cytotoxic effector molecule expression. These immunological differences open up the possibility of different disease aetiology and pathogenesis mechanisms in the two groups. These findings may suggest that people with ME-MM have frequent antigen exposure, likely related to persistent viral infection and frequent reactivation, leading to the appearance of early senescence memory T cells and MAIT cells, and also the expression of more cytotoxic effector molecules in cytotoxic cells with diminished CD8 molecules on CD8^+^T cells. In contrast, people with ME-SA had an ongoing uncontrolled pro-inflammatory immune system activation, with more activated T cells in blood, and higher cell activation and pro-inflammatory cytokine production in response to stimulation. The sustained activation and inflammatory cytokine production in ME-SA may be a cause or result of symptom exacerbation, and may contribute to symptom severity. These differences support the importance of careful patient stratification in the clinical management of ME/CFS, and also in the development of new diagnostic tools and treatments.

## Methods

### Sex as a biological variable

Both female and male participants were included in the study. ME/CFS predominantly affects women, and our study cohort of 96 individuals included 76 women (79%) and 20 men (21%) which reflects this difference in susceptibility.

### Study Participants

A total of 96 participants were enrolled in the study. All participants with ME/CFS had a formal diagnosis and met either the Canadian Consensus Criteria (9) or CDC-1994 criteria (31); many fulfil both (30). Symptom questionnaires and clinical appointments were used to assess the severity of disease, with participants characterised as having mild/moderate ME/CFS (ME-MM) or being severely affected (ME-SA) (8). Severely affected study participants were visited at home by the clinical team. The study was designed as a longitudinal study, with blood samples collected five times with at least six month gaps between time points. However, the Covid-19 pandemic negatively affected follow-up, so participants were variously sampled 2, 3, 4 or 5 times. The participants’ demographic information is shown in Table 1. Venous blood from participants was collected into heparinized vacutainers, and transferred to the UCL/RFH Biobank at the Royal Free Hospital London for isolation and cryopreservation of peripheral blood mononuclear cells (PBMCs).

### Flow cytometry

Cryopreserved PBMCs were transferred to the London School of Hygiene & Tropical Medicine (LSHTM) for analysis, where they were stored in liquid nitrogen until use. On the experiment day, PBMC vials were defrosted in a 37°C water bath for 1 min, then the contents transferred to 12 ml warm RPMI 1640 including 10 % foetal bovine serum (RPMI/FCS) and centrifuged at 650 g for 15 mins. After 2 washes with RPMI including 10 % FBS, cells were rested at 37°C/5% CO_2_ for 30 mins. Cells were then pelleted by centrifugation and re-suspended in RPMI/FCS, after which cells were counted and viability ascertained using the Countess3 automated cell counter (Invitrogen, MA, USA). One million cells were added to wells of a 96 well V-bottom plate for “ex vivo” staining, where extra- and intra-cellular staining processes with target antibodies were performed: antibodies used are shown in Supplemental Table 1. Another one million cells/sample were incubated overnight in RPMI/FCS at 37°C/5% CO_2_ in preparation for “*In vitro* stimulation” the next day. A total of five staining panels were used for flow cytometric analysis of each sample, including four ex vivo and one in vitro panel (Supplemental Table 2).

For extracellular staining, cell suspensions were centrifuged at 750 g for 5 mins, supernatants were removed by flicking, and cell pellets were loosened by gentle vortex. Cells were washed with FACS buffer containing bovine calf serum and sodium azide (Cell Staining Buffer, BioLegend, CA, USA) and centrifuged. Cells were first incubated with 1:200 prediluted MR1 tetramer in 20 μl FACS buffer (loaded with 5-OP-RU and 6-FP as a negative control) obtained from the NIH Tetramer Core Facility (32), for 40 mins at room temperature in the dark. Cells were then washed and stained with the appropriate cocktail of antibodies and live/dead stain in FACS buffer at 4 °C for 20 min: these antibody staining panels included anti-human CD3/CD4/CD8/CD56/CD161/TCR Vα7.2 for T cell, MAIT and NK cell phenotyping throughout, along with “activation/exhaustion” and “memory/differentiation” specific marker staining panels (Supplemental Table 2). After incubation, 200μl of FACS buffer was added, and cells pelleted by centrifugation.

For the “function” panel, for both *ex vivo* and *in vitro* stimulated cells, following the extracellular staining, 75μl of cell fixation and permeabilization solution (BD biosciences, NJ, USA) was added to each well, and incubated for 15 mins at room temperature in the dark. Then, using ‘Perm/Wash buffer (BD biosciences), cells were washed and stained with anti-cytokines/cytotoxic granule antibodies for 30 mins at room temperature in the dark. Similarly, for the “Transcription factor” panel, cells were fixed and permeabilized using the FOXP3 Transcription factor staining buffer set (eBioscience, CA, USA) for 30 mins at room temperature in the dark, then stained with anti-human transcription factor antibodies for 1 hour at room temperature in the dark, in permeabilization buffer. After intracellular or intranuclear staining, cells were centrifuged at 930 g for 5 mins, then suspended with FACS buffer, and analysed by flow cytometry the following day.

For the *in vitro* stimulation, following the overnight incubation, “cell stimulation cocktail” (phorbol 12-myristate 13-acetate (PMA) and ionomycin, eBioscience) was added to the cells in the 24-well plate. After 1 hour, protein transport inhibitor cocktail (brefeldin A and monensin, eBioscience) was added to each well, then incubated for 4 hours more. After incubation, the supernatant was removed by pipetting, and all cells were transferred to a 96 well V-bottom plate, and stained as described above.

To validate the multiparametric flow cytometry panels, they were first optimised on samples from healthy control donors. To monitor the consistency of flow cytometry across different batches of experiments, we included PBMCs from healthy controls in all experiments. FMOs (fluorescence minus one) and isotype controls were included for the gating strategy in all experiments.

An LSRII cytometer (BD) was used for cell acquisition, and flow cytometry data analysis was done using FlowJo software version 10.8.1 (Tree star Inc. Ashland, USA).

### Data Availability

The data underlying the analysis can be accessed in the Supporting Data Values Excel file.

### Statistics

Presented values (cell frequency or Median Fluorescence Intensity (MFI)) were calculated as the mean value of those acquired at different timepoints (2,3,4,5). Datasets were tested for normality using the Shapiro-Wilk test. For two group comparisons, the t-test or Mann-Whitney U-test were used for normal or non-normal distribution data respectively. For paired sample comparisons, the paired t-test for parametric data or Wilcoxon matched-pairs test for non-parametric data were used. Spearman correlation tests were conducted to determine the association between two variables. A P value <0.05 was considered statistically significant, and tests were 2-sided. All flow cytometry data analyses were performed using GraphPad Prism version 10.0.2.

### Study Approval

This study was conducted according to the principles of the Declaration of Helsinki. Study approval was obtained from the LSHTM Medicine Research Ethics Committee 16 January 2012 (Ref.6123) and the National Research Ethics Service (NRES) London: Bloomsbury Research Ethics Committee 22 December 2011 (REC ref.11/10/1760, IRAS ID: 77765). Written informed consent was obtained from all study participants prior to inclusion in the study.

## Supporting information

Supporting Data Values

## Author Contributions

LN, JMC, HD designed the research studies; EL, CCK, JLS, GS conducted the experiments; JLS, GS acquired the data; JLS, EL, JMC analysed the data; JLS, JMC wrote the manuscript; All authors reviewed the manuscript.

## Acknowledgements

We thank all the study participants, for donating their blood to the UK ME/CFS Biobank and for giving us their time and their energy. We are grateful to Dr Martin Goodier for flow cytometry advice. The analysis was mainly funded by the NIH: R01AI103629, with additional support from NIH: 5R01AI170839-03 and from The ME Association.

## Notes

### Competing Interest Statement

The authors have declared no competing interest.

