## Supplemental Tables for "Abnormal T-Cell Activation And Cytotoxic T-Cell Frequency Discriminates Symptom Severity In Myalgic Encephalomyelitis/Chronic Fatigue Syndrome"

Supplementary Table S1: Fluorescently labelled antibodies used for flow cytometry.

| | | Fluorochrome | manufacturer | Clone | Isotype |
|---|---|---|---|---|---|
| Cell phenotyping | CD3 | AF700 | ebioscience | UCHT1 | Mouse IgG1, κ |
| | CD4 | V500 | BD Horizon | RPA-T4 | Mouse IgG1, κ |
| | CD8 | BV711 | BioLegend | RPA-T8 | Mouse IgG1, κ |
| | CD56 | BV650 | BioLegend | HCD56 | Mouse IgG1, κ |
| | MR1 5-OP-RU tetramer | PE | Gifted from NIH | | |
| | CD161 | PerCP-Cy5.5 | ebioscience | HP-3G10 | Mouse IgG1, κ |
| | TCR Vα7.2 | BV605 | BioLegend | 3C10 | Mouse IgG1, κ |
| | Live/dead/ Near-IR | APC/Cy7 | ebioscience | | |
| Naïve/Memory /differentiation | CCR7 | APC | Biolegend | G043H7 | Mouse IgG2a, κ |
| | CD45RA | BB515 | BD Horizon™ | HI100 | Mouse IgG2b, κ |
| | CD28 | PerCP-Cy5.5 | Biolegend | CD28.2 | Mouse IgG1, κ |
| | CD57 | eFluor 450 | eBioscience | TB01 | Mouse IgM |
| Activation /Exhaustion | PD-1 | FITC | BioLegend | EH12.2H7 | Mouse IgG1, κ |
| | CD69 | APC | BioLegend | FN50 | Mouse IgG1, κ |
| | CD38 | PE-eFluor 610 | eBioscience | HIT2 | Mouse IgG1, κ |
| | TIM-3 | BV421 | BioLegend | F38-2E2 | Mouse IgG1, κ |
| Transcription Factor | PLZF | AF488 | eBioscience | Mags.21F7 | Mouse IgG1, κ |
| | T-bet | eFlour660 | eBioscience | eBio4B10 (4B10) | Mouse IgG1, κ |
| | EOMES | PE-eFluor 610 | eBioscience | WD1928 | Mouse IgG1, κ |
| | RORgt | BV421 | BD Biosciences | Q21-559 | Mouse IgG2b, κ |
| Function (cytokine/ cytotoxic molecules) | GrzB | FITC | BioLegend | GB11 | Mouse IgG1, κ |
| | IL17A | eFlour660 | eBioscience | eBio64CAP17 | Mouse IgG1, κ |
| | IFNg | PE-eFluor 610 | eBioscience | 4S.B3 | Mouse IgG1, κ |
| | Prf | BV421 | BioLegend | B-D48 | Mouse IgG1, κ |
| | TNFa | eFluor 450 | eBioscience | MAb11 | Mouse IgG1, κ |
| | PD-1 | BV650 | BioLegend | EH12.2H7 | Mouse IgG1, κ |
| | CD69 | PE-Cy7 | eBioscience | FN50 | Mouse IgG1, κ |

**Supplementary Table S2:** Panel used flow cytometry analyses.

| | *Ex vivo* | | | | | *In vitro* |
|---|---|---|---|---|---|---|
| | Panel 1 | Panel 2 | Panel 3 | Panel 4 | Panel 5 | Panel 6 |
| Fluorochromes | Activation/ Exhaustion | Memory/differentiation | Function | Transcription Factor | Combined panels 1 to 4 | Function |
| AF700 | CD3 | CD3 | CD3 | CD3 | CD3 | CD3 |
| V500 or BV510 | CD4 | CD4 | CD4 | CD4 | CD38 | CD4 |
| BV711 | CD8 | CD8 | CD8 | CD8 | Perforin | CD8 |
| BV650 | CD56 | CD56 | CD56 | CD56 | PD-1 | PD-1 |
| BV605 | TCR Vα7.2 | TCR Vα7.2 | TCR Vα7.2 | TCR Vα7.2 | T-bet | TCR Vα7.2 |
| PerCP-Cy5.5 | CD161 | CD28 | CD161 | CD161 | GranzymeB | CD161 |
| PE | MR1 5-OP-RU+ | MR1 | MR1 | MR1 | CD4 | MR1 |
| Fixable Near-IR | Viability | Viability | Viability | Viability | Viability | Viability |
| FITC or BB515 or AF488 | PD-1 | CD45RA | GranzymeB | PLZF | CD45RA | GranzymeB |
| APC or Ef660 | CD69 | CCR7 | IL-17 | T-bet | CCR7 | IL-17 |
| PE-eFluor610 | CD38 | | IFNγ | EOMES | EOMES | IFNγ |
| BV421 or eFlour450 | TIM-3 | CD57 | Perforin | RORγt | HLA-DR | TNFα |
| PE-Cy7 | | | | | CD8 | CD69 |
