## Supplemental Figures for "Abnormal T-Cell Activation And Cytotoxic T-Cell Frequency Discriminates Symptom Severity In Myalgic Encephalomyelitis/Chronic Fatigue Syndrome"

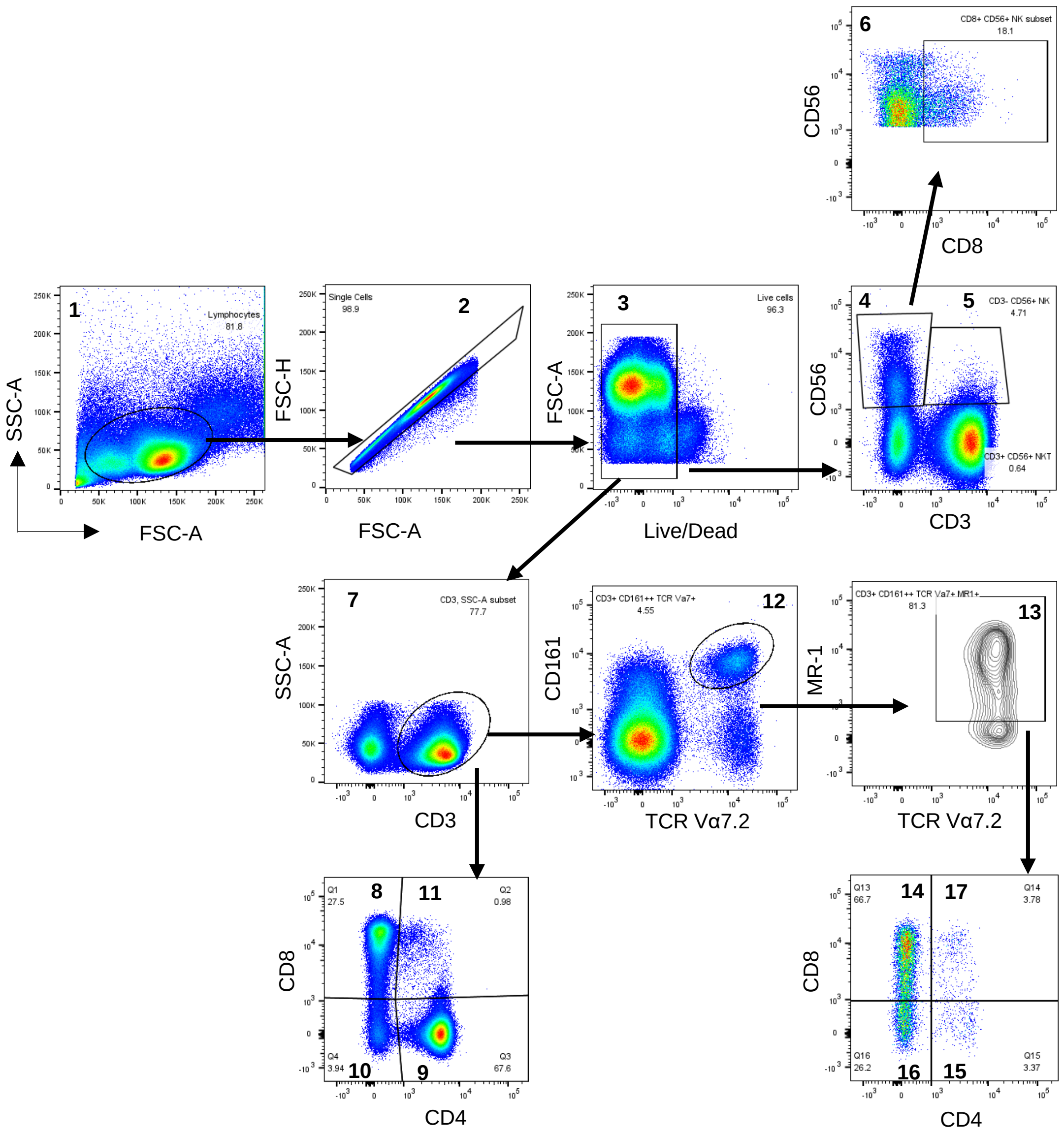


**Supplemental Figure 1: Lymphocyte gating strategy for phenotyping of PBMC.** Thawed PBMC from people with ME/CFS were stained with fluorescently labelled antibodies (Supplementary Table S1). Data were collected by flow cytometry and analysed using FlowJo software. (1) Lymphocytes were identified by forward and side scatter area profile, (2) singlet cells were identified by forward scatter height and area, and the other cell types were identified as positive populations based on gating by FMO controls or isotype controls. (1) lymphocytes, (2) singlet, (3) live cells, (4) NK cells as CD3-CD56+, (5) NKT-like cells as CD3+CD56+, (6) NK8 cells as CD3-CD56+CD8+, (7) T cells as CD3+. Within the T cell gate, T cells were characterised further based on CD4 and CD8 expression: (8) CD8+T cells, (9) CD4+T cells, (10) CD4-CD8- double negative (DN) cells, (11) CD4+CD8+ double positive (DP) cells. In parallel, The T cell gate was characterised by expression of TCR Vα7.2 and CD161 expression. (12) CD3+TCR Vα7.2+CD161++ cells (generally recognised as MAIT cells), (13) In this study, MAIT cells were defined as CD3+TCR Vα7.2+CD161++MR-1+. Further, the MAIT cells were characterised based on expression of CD4 and CD8: (14) CD8+ MAIT cells, (15) CD4+ MAIT cells, (16) CD4-CD8- DN MAIT cells, (17) CD4+CD8+ DP MAIT cells. This figure was a representative example obtained using the 'activation/exhaustion' staining panel.

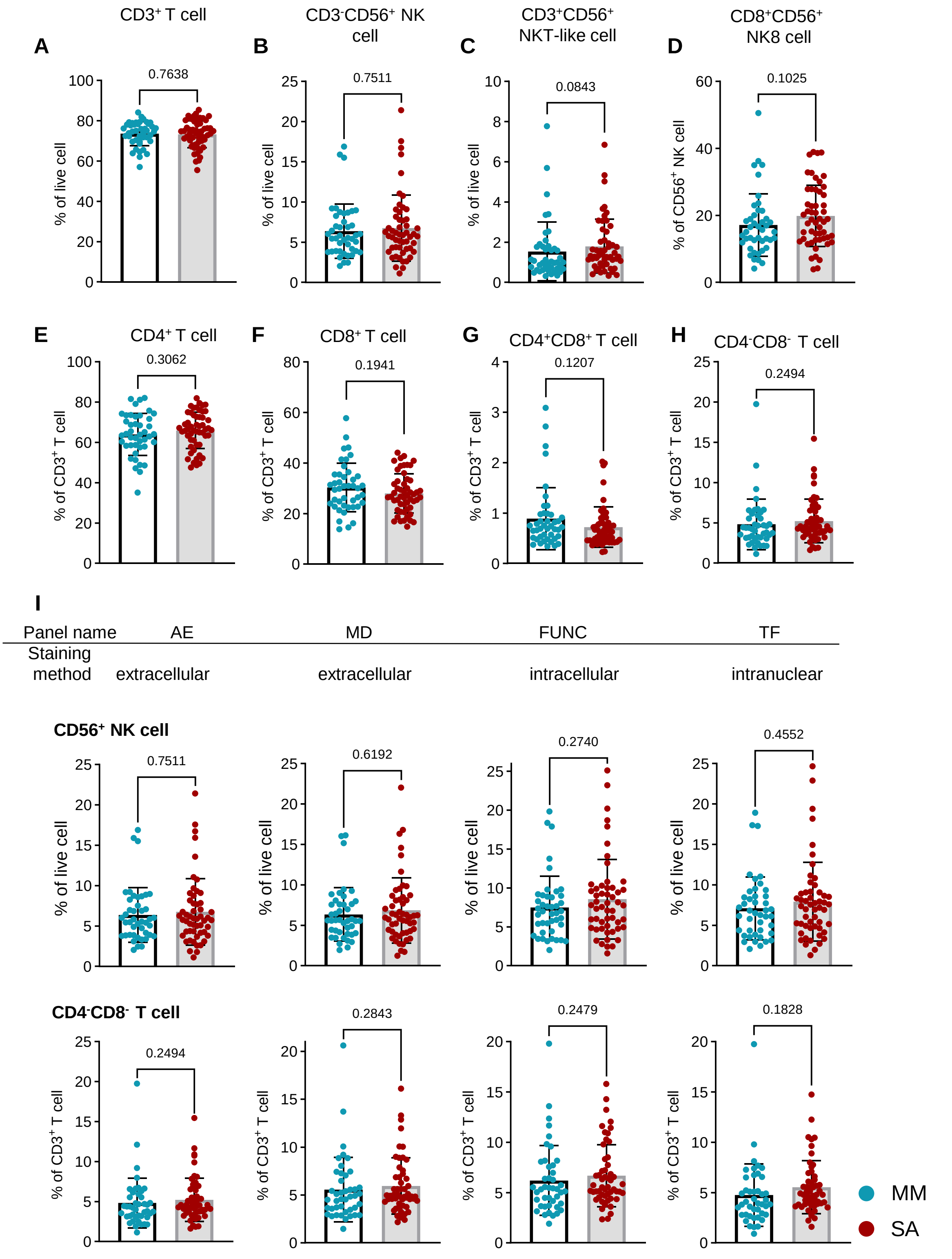


**Supplemental Figure 2: Comparison of frequencies of T cells and NK cells and their subsets in people with mild/moderate (n=43) and severe ME/CFS (n=53).** The frequencies of (A) T cells, (B) NK cells, (C) NKT-like cells, (D) NK8 cells, (E) $CD4^{+}$T cells, (F) $CD8^{+}$T cells, (G) $CD4^{-}CD8^{-}$T cells, and (H) $CD4^{+}CD8^{+}$T cells were compared between the two clinical groups, using data obtained from the 'activation/exhaustion marker' staining panel. (I) Frequencies of NK cells and $CD4^{-}CD8^{-}$ DN T cells in *ex vivo* PBMC stained by using different panels are shown. "AE": Activation/Exhaustion markers; ''MD'': Memory/Differentiation; "Func": functional markers; "TF": transcription factor markers. Each dot represents the average value across all the samples collected at different time points for individual study participants. Mean values and SD for each group are shown. Data were compared using the Mann-Whitney test for non-parametric data, with $p<0.05$ deemed significant. MM: people with mild/moderate symptoms, SA; severely affected people.

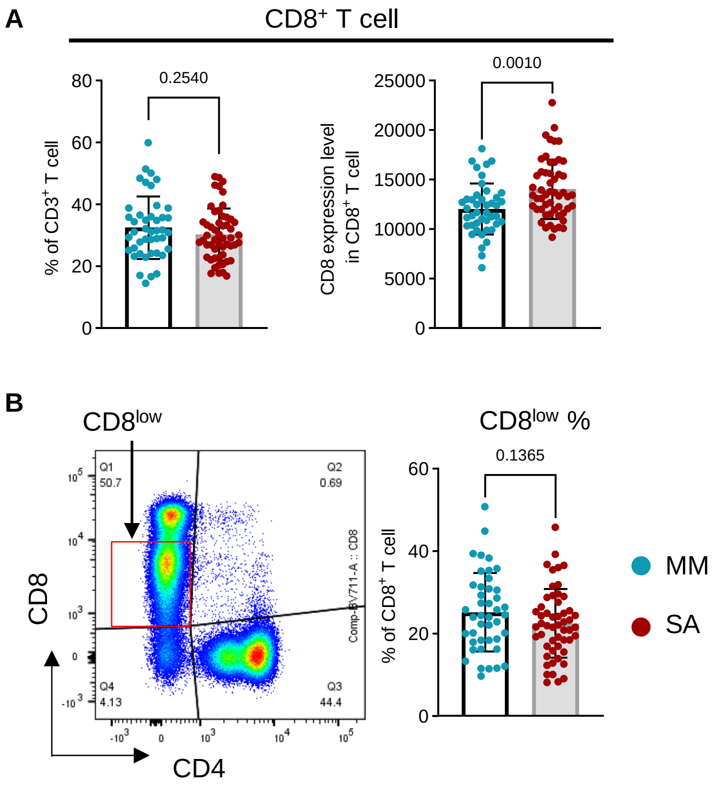


**Supplemental Figure 3: Differential median fluorescence intensity of CD8α in people with mild/moderate (n=43) and severe ME/CFS (n=53).** (A) The frequency of CD8$^{+}$ T cells (left) and median fluorescence intensity of CD8α (right) were compared between the two groups. The dataset was from 'functional marker' staining panel. (B) Gating for CD8$^{low}$ population is shown as a representative pseudocolour plot (left) and the CD8$^{low}$ population percentage in the CD8$^{+}$T cells was compared between two groups (right). Each dot represents the average value across all the samples collected at different time points for individual study participants. Mean values and SD are shown. Datasets were compared using the Mann-Whitney test for non-parametric data, with $p<0.05$ deemed significant. MM: people with mild/moderate symptoms; SA: severely affected people.

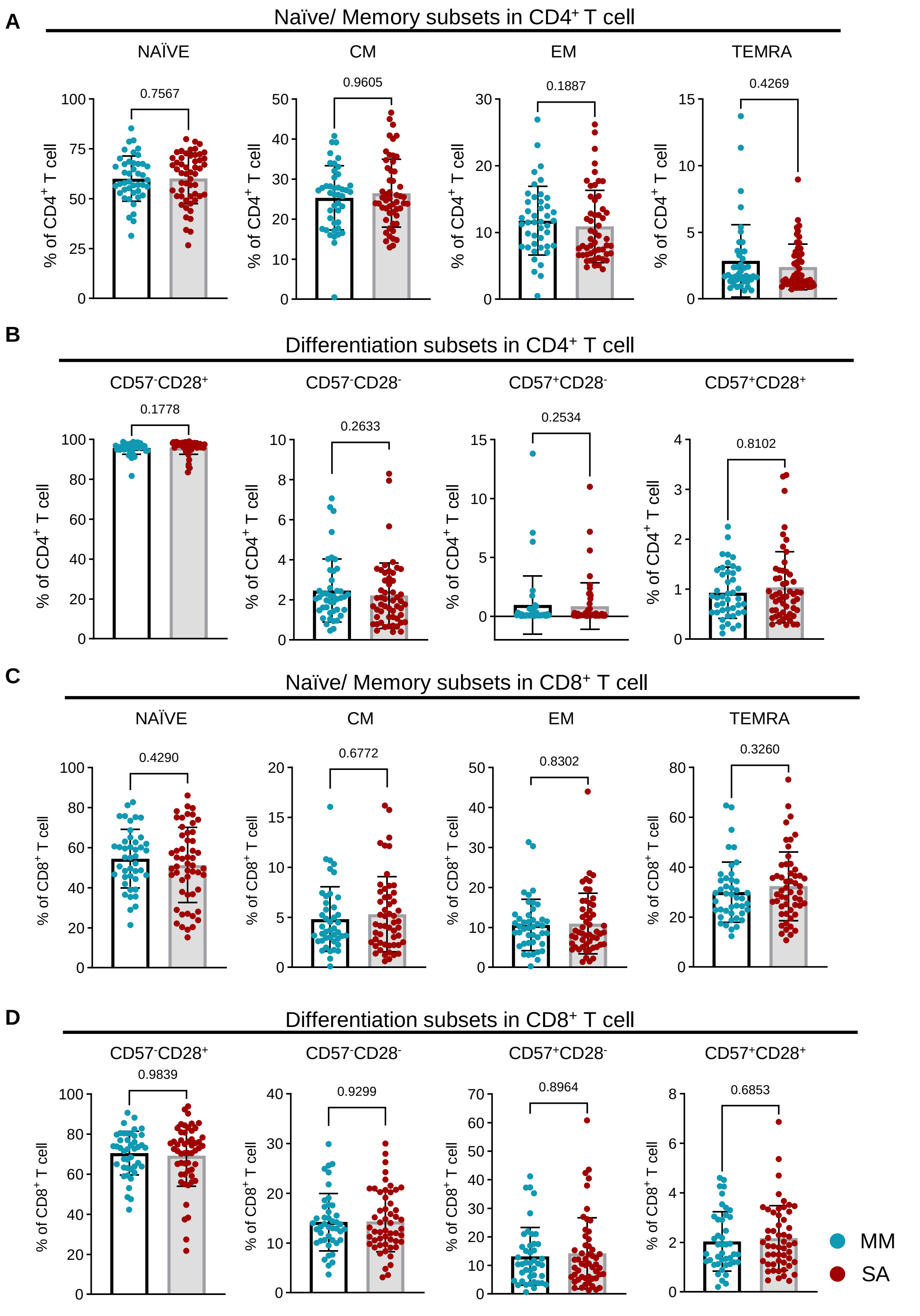


**Supplemental Figure 4: Frequencies of naïve/memory and differentiation subsets in $CD4^+$ T cells and $CD8^+$ T cells from people with mild/moderate (n=43) and severe ME/CFS (n=53).** *Ex vivo* PBMC were analysed using the ''memory/differentiation panel'', comprised of CD45RA, CCR7, CD57, CD28. Within the $CD3^+$T cell compartment, $CD4^+$ and $CD8^+$T cells were analysed by expression of CCR7 and CD45RA to define naïve and memory cells (A and C) and CD57 and CD28 to define differentiation status (B and D). The frequencies were compared between the two clinical groups. Naïve: $CCR7^+CD45RA^+$; Central Memory (CM): $CCR7^+CD45RA^-$; Effector Memory (EM):$CCR7^-CD45RA^-$; terminally re-expressing CD45RA effector cells (TEMRA): $CCR7^-CD45RA^+$. Each dot represents the average value across all the samples collected at different time points for individual study participants. Mean values and SD are shown. Datasets were compared using the Mann-Whitney test for non-parametric data, with $p<0.05$ deemed significant. MM: people with mild/moderate symptoms; SA: severely affected people.

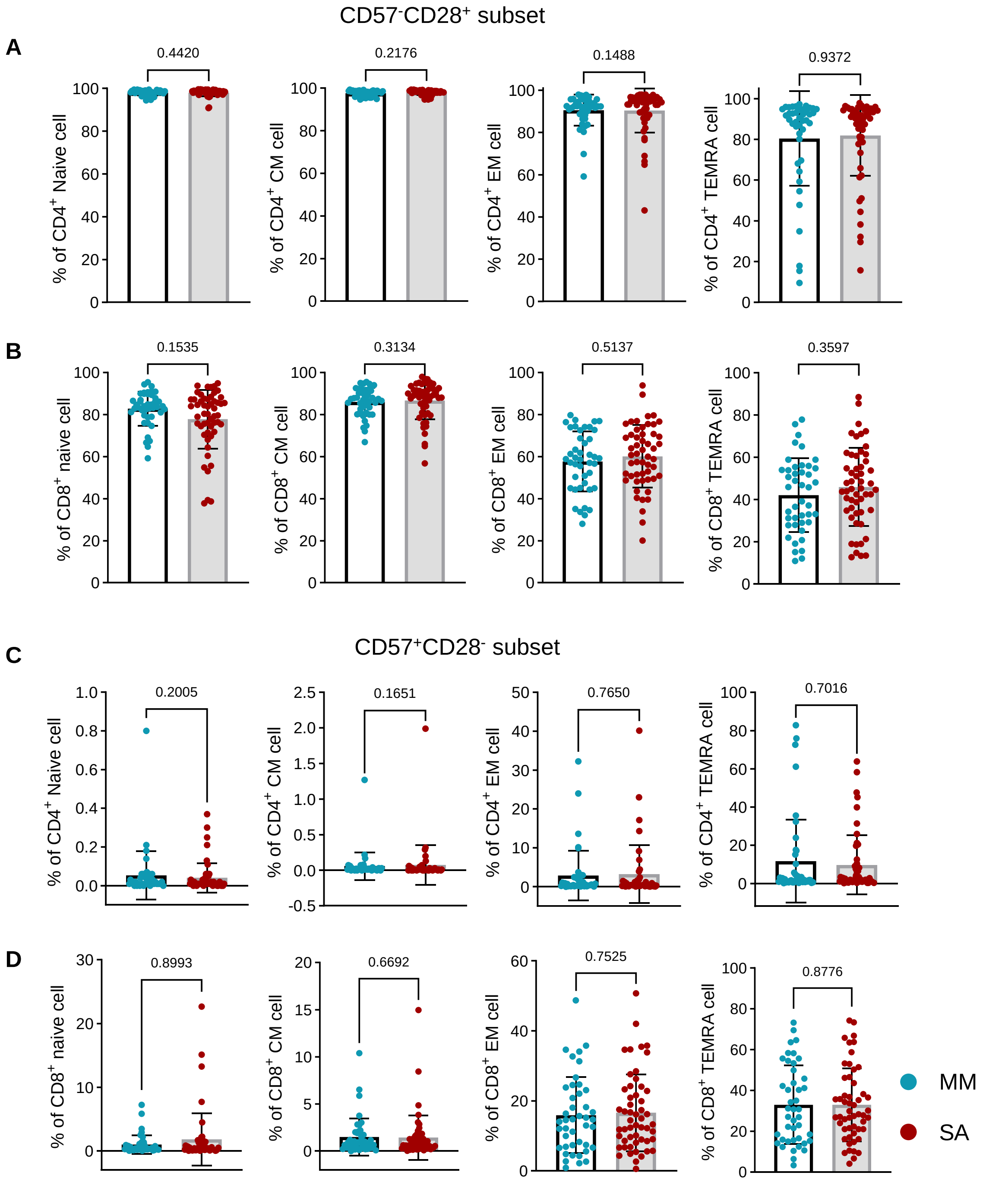


**Supplemental Figure 5: Analysis of naïve /memory subsets of CD4$^+$ and CD8$^+$ T cells by expression of CD57 and CD28 in people with mild/moderate (n=43) and severe ME/CFS (n=53).** *Ex vivo* PBMC were analysed using the 'memory/differentiation' staining panel, comprised of CD45RA, CCR7, CD57, and CD28. Each CD4$^+$ and CD8$^+$ naïve/memory T cell subset was further analysed for expression of CD57 and CD28, to define CD57$^-$CD28$^+$, CD57$^-$CD28$^-$, CD57$^+$CD28$^-$ and CD57$^+$CD28$^+$ subsets. The frequencies of the CD57$^-$CD28$^+$ subset (A, B) and the CD57$^+$CD28$^-$ subset (C, D) are shown for naïve, CM, EM and TEMRA CD4$^+$ (A, C) and CD8$^+$ (B, D) T cells. Each dot represents the average value across all the samples collected at different time points for individual study participants. Mean values and SD are shown, and datasets were compared using the Mann-Whitney test for non-parametric data or the t-test for parametric data, with $p<0.05$ deemed significant. MM: people with mild/moderate symptoms; SA: severely affected people.

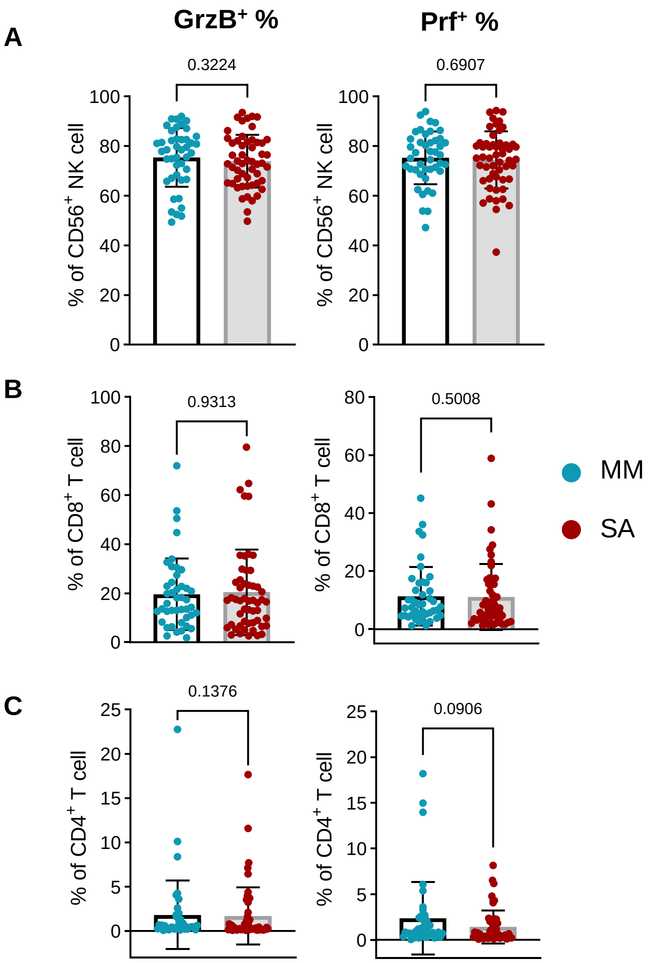


**Supplemental Figure 6: Comparison of frequencies of cytotoxic mediators (granzyme B and perforin) in NK cells, CD8⁺ T cells and CD4⁺ T cells from people with mild/moderate (n=43) and severe ME/CFS (n=53).** *Ex vivo* PBMC were stained with fluorescently labelled antibodies extracellularly for immune cell phenotyping, then stained with antibodies from the 'functional panel', comprised of granzyme B, perforin, IL-17 and Interferon-γ intracellularly. The frequencies of granzyme B and perforin were compared between the two groups in NK cells (A), $CD8^+$ T cells (B) and $CD4^+$ T cells (C). Each dot represents the average value across all the samples collected at different time points for individual study participants. Mean values and SD are shown. Datasets were compared using the Mann-Whitney test for non-parametric data or the t-test for parametric data, with $p<0.05$ deemed significant. MM: people with mild/moderate symptoms; SA: severely affected people. GrzB; granzyme B, Prf; perforin.

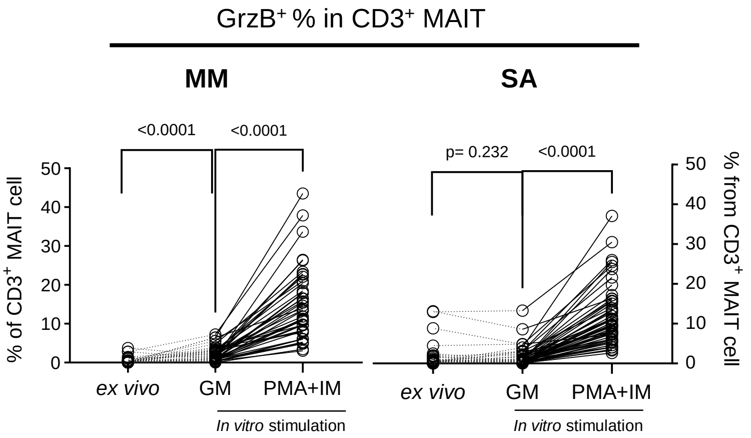


**Supplemental Figure 7: Increased frequencies of granzyme B-expressing MAITs in PBMCs following overnight incubation from people with mild/moderate ME/CFS.** Cells were analysed on the day of cell thawing (‘*ex vivo*’), or the following day without stimulation (“GM”), or following overnight incubation and then 5 hours of stimulation with PMA and ionomycin (“PMA+IM”). PBMCs were stained extracellularly with antibodies for MAIT cell phenotyping, then intracellularly from the ‘functional panel’, comprised of granzyme B, perforin, IL-17 and Interferon-γ antibodies. The frequencies of granzyme B were compared between different conditions in people with ME-MM (left) and ME-SA (right). Each dot represents the average value across all the samples collected at different time points for individual study participants. For paired sample comparison (ex vivo vs GM or GM vs PMA+IM), the Wilcoxon matched-pairs test was used for non-parametric data, with $p<0.05$ deemed significant. MM: people with mild/moderate symptoms; SA: severely affected people. GrzB; granzyme B, GM; growth medium, PMA; phorbol 12-myristate 13-acetate, IM; ionomycin.

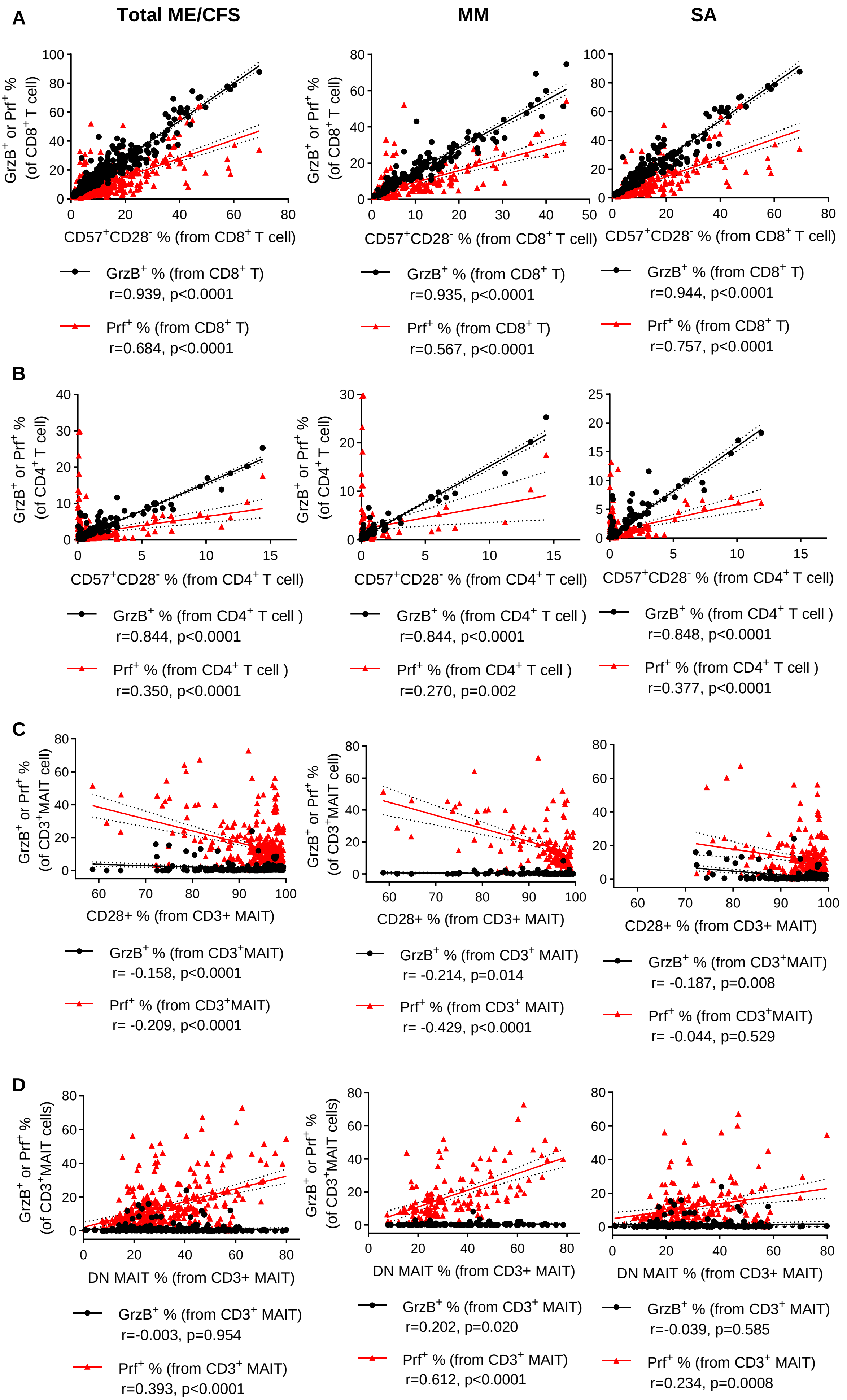

A
Total ME/CFS
MM
SA
GrzB+ or Prf+ % (of CD8+ T cell)
CD57+CD28- % (from CD8+ T cell)
GrzB+ % (from CD8+ T)
r=0.939, p<0.0001
Prf+ % (from CD8+ T)
r=0.684, p<0.0001
GrzB+ % (from CD8+ T)
r=0.935, p<0.0001
Prf+ % (from CD8+ T)
r=0.567, p<0.0001
GrzB+ % (from CD8+ T)
r=0.944, p<0.0001
Prf+ % (from CD8+ T)
r=0.757, p<0.0001
B
GrzB+ or Prf+ % (of CD4+ T cell)
CD57+CD28- % (from CD4+ T cell)
GrzB+ % (from CD4+ T cell )
r=0.844, p<0.0001
Prf+ % (from CD4+ T cell )
r=0.350, p<0.0001
GrzB+ % (from CD4+ T cell )
r=0.844, p<0.0001
Prf+ % (from CD4+ T cell )
r=0.270, p=0.002
GrzB+ % (from CD4+ T cell )
r=0.848, p<0.0001
Prf+ % (from CD4+ T cell )
r=0.377, p<0.0001
C
GrzB+ or Prf+ % (of CD3+MAIT cell)
CD28+ % (from CD3+ MAIT)
GrzB+ % (from CD3+MAIT)
r= -0.158, p<0.0001
Prf+ % (from CD3+MAIT)
r= -0.209, p<0.0001
GrzB+ % (from CD3+ MAIT)
r= -0.214, p=0.014
Prf+ % (from CD3+ MAIT)
r= -0.429, p<0.0001
GrzB+ % (from CD3+MAIT)
r= -0.187, p=0.008
Prf+ % (from CD3+MAIT)
r= -0.044, p=0.529
D
GrzB+ or Prf+ % (of CD3+MAIT cells)
DN MAIT % (from CD3+ MAIT)
GrzB+ % (from CD3+ MAIT)
r=-0.003, p=0.954
Prf+ % (from CD3+ MAIT)
r=0.393, p<0.0001
GrzB+ % (from CD3+ MAIT)
r=0.202, p=0.020
Prf+ % (from CD3+ MAIT)
r=0.612, p<0.0001
GrzB+ % (from CD3+ MAIT)
r=-0.039, p=0.585
Prf+ % (from CD3+ MAIT)
r=0.234, p=0.0008

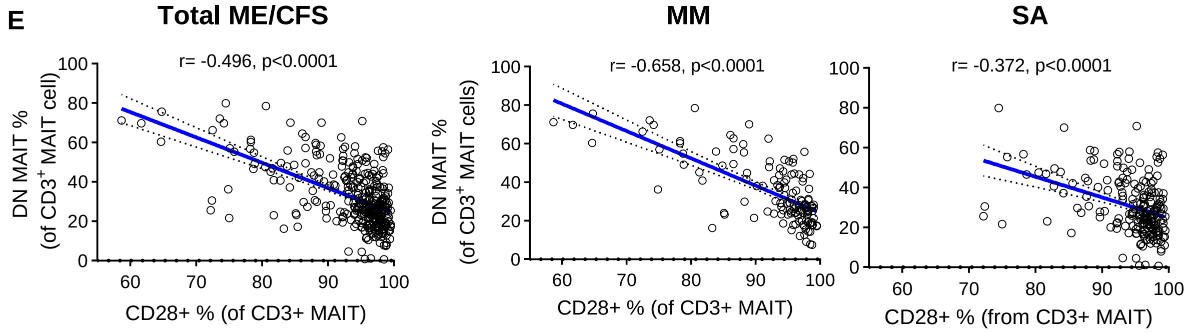


**Supplemental Figure 8: Correlation analysis between frequencies of cytotoxic T cell subsets and other T cell subsets with different differentiation status in the total sample set (n=335) or by two clinical groups separately (n=132 for ME-MM, n=203 for ME-SA).** For this analysis, the data collected from ‘memory/differentiation’ and ‘function’ flow cytometry staining panels were used. Spearman Correlation analyses were performed between frequencies of A) cytotoxic molecule-positive $CD8^{+}$T cells and $CD57^{+}CD28^{-}CD8^{+}$T cells, B) cytotoxic molecule-positive $CD4^{+}$T cells and $CD57^{+}CD28^{-}CD4^{+}$T cell subsets, C) cytotoxic molecule-positive MAITs and $CD28^{+}$ MAITs, D) cytotoxic molecule-positive MAITs and DN MAITs, and E) CD28+MAIT cells and DN MAIT cells. Dashed lines represent the 95% confident intervals. Each dot or triangle represents the average value across all the samples collected at different time points for individual study participants. p-value<0.05 is deemed significant. MM: people with mild/moderate ME/CFS; symptoms, SA: people severely affected with ME/CFS.

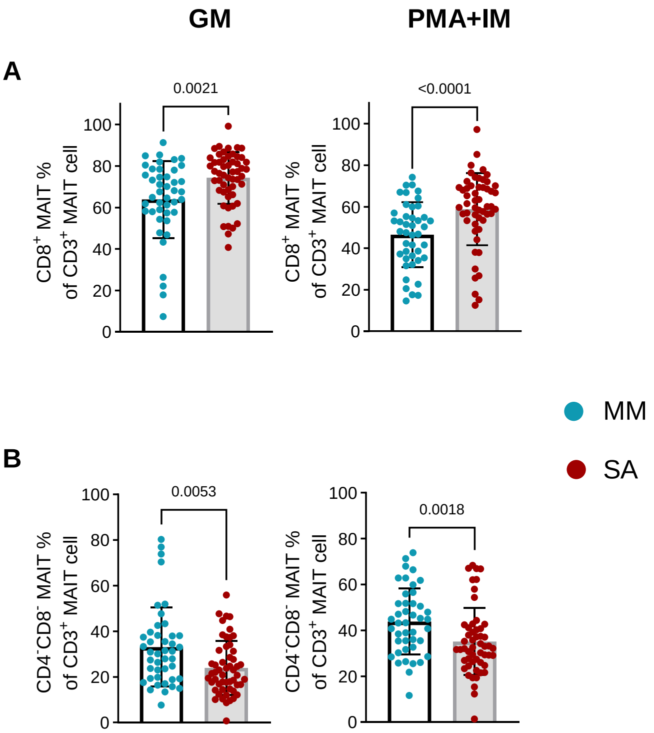


**Supplemental Figure 9: Proportion changes of MAIT cell subsets after stimulation with PMA and ionomycin in people with mild/moderate (n=43) and severe ME/CFS (n=53).** Thawed PBMCs were incubated overnight, and then stimulated with PMA and ionomycin for 5 hours. Then the stimulated cells were stained for surface T cell markers (CD4, CD8, MAIT). The unstimulated cells were stained in parallel. Each dot represents the average value across all the samples collected at different time points for individual study participants. Mean values and SD are shown. Datasets were compared using the Mann-Whitney test for non-parametric data or the t-test for parametric data, with p<0.05 deemed significant. MM: people with mild/moderate symptoms; SA: severely affected people. GM; growth medium, PMA; Phorbol 12-myristate 13-acetate, IM; ionomycin

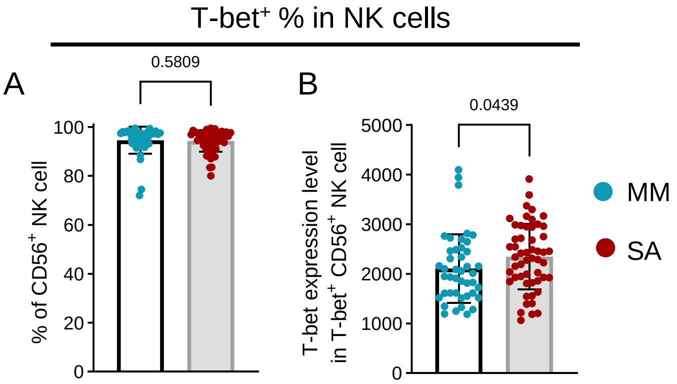


**Supplemental Figure 10: Elevated median fluorescence of intensity (MFI) of T-bet expressing NK cells in people with severe ME/CFS.** *Ex vivo* PBMC were stained for immune cell phenotyping, then intranuclearly stained with antibodies from the 'transcription factor panel'. The frequency of T-bet expressing NK cells and MFI of T-bet were compared between the two clinical groups (A: frequency of T-bet$^{+}$ population in NK cells; B: MFI of T-bet expression in T-bet$^{+}$ NK cells). Each dot represents the average value across all the samples collected at different time points for individual study participants. Mean values and SD are shown. Datasets were compared using the Mann-Whitney test for non-parametric data, with $p<0.05$ deemed significant. MM: people with mild/moderate ME/CFS; SA: people severely affected by ME/CFS.

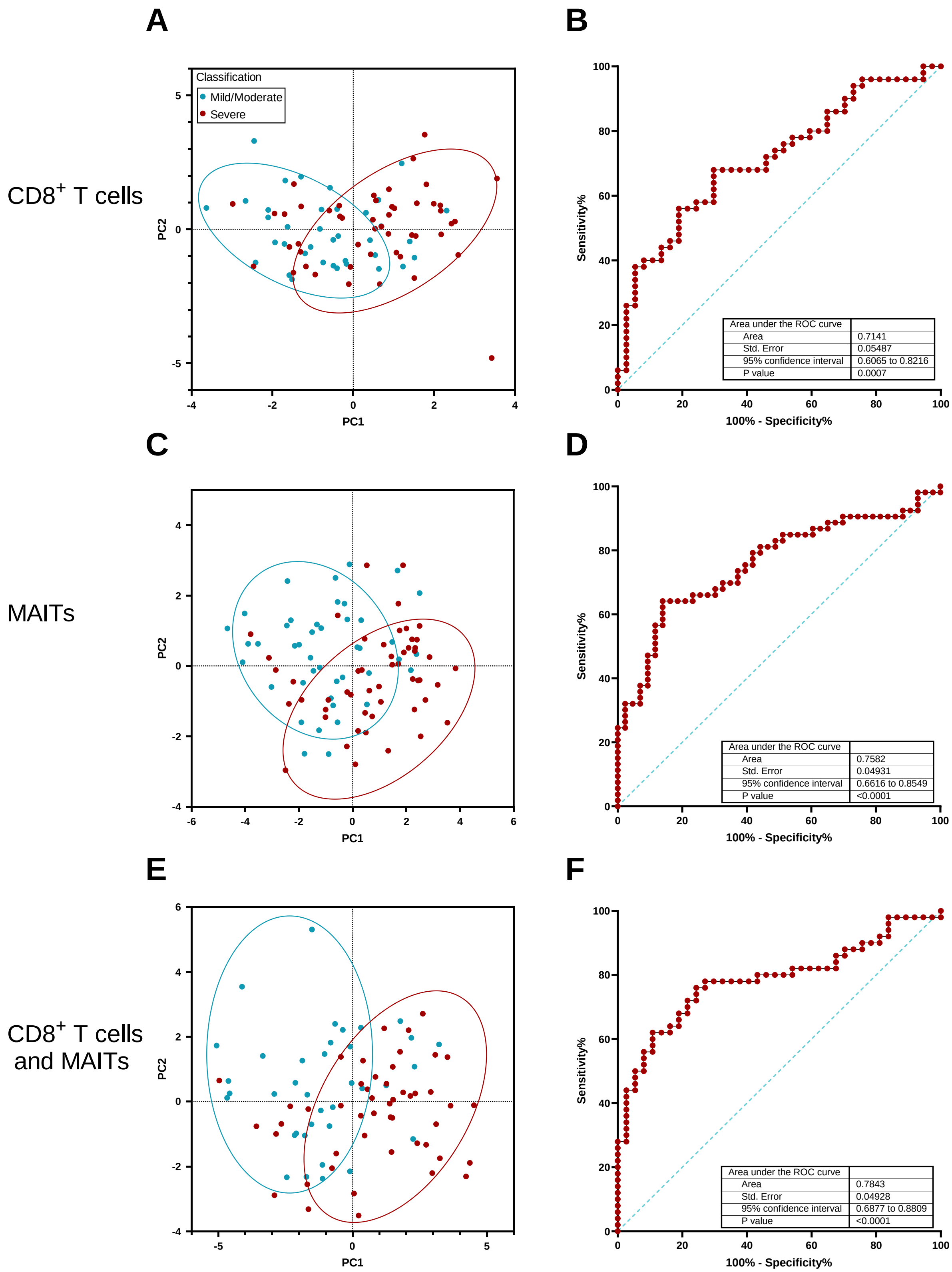


**Supplemental Figure 11: Discrimination between people with mild/moderate or severe ME/CFS based on differential expression of markers in CD8$^+$ T cells and MAITs.** PCA plots of ME-MM and ME-SA study participants, based on the expression of subsets of markers in CD8$^+$ T cells (A), MAITs (C) or both CD8$^+$ and MAITs (E). Receiver Operating Characteristic curve analysis of ratios of activation / senescence-cytotoxicity markers in ME-SA vs ME-MM study participants in B) CD8$^+$ T cells ((evCD69$^+$ + ivIFNγ$^+$) / (CD28$^-$CD57$^-$CD8$^+_{EM}$ + CD28$^-$CD57$^-$CD8$^+_{TEMRA}$)), D) MAITs ((%CD8$^+$MAITs + ivTNF$^+$) / (evPrf$^+$ + ivGzB$^+$)) or F) both CD8$^+$ and MAITs (((evCD69$^+$ + ivIFNγ$^+$) / (CD28$^-$CD57$^-$CD8$^+_{EM}$ + CD28$^-$CD57$^-$CD8$^+_{TEMRA}$)) x ((%CD8$^+$MAITs + ivTNF$^+$) / (evPrf$^+$ + ivGzB$^+$)). Parameters used in PCA analyses: CD8$^+$ T cells (A & E): %CD28$^-$CD57$^-$CD8$^+_{EM}$, %CD28$^-$CD57$^-$CD8$^+_{TEMRA}$, MFI-Prf evCD8$^+$, MFI-GzB evCD8$^+$, MFI-PD1 evCD8$^+$, %evCD69$^+$CD8$^+$, %evCD69$^+$CD38$^+$CD8$^+$, % ivIL-17+ CD8$^+$, %ivTNF$^+$CD8$^+$, %ivIFNγ$^+$ CD8$^+$; MAITs (C & E): %MAITs, %CD8$^+$MAITs, %evCD69$^+$MAITs, %evCD69$^+$CD38$^+$MAITs, %ivCD69$^+$MAITs, %ivTNF$^+$MAITs %ivIFNγ$^+$MAITs, %ivTNF$^-$IFNγ$^-$MAITs, %evT-bet$^+$MAITs, %evCD28$^+$MAITs, %ivGzB$^+$MAITs, %evPrf$^+$MAITs, MFI-PD1 evMAITs. Rings in the PCA plots contain 80% of the corresponding participants. ev: *ex vivo*; iv: following stimulation *in vitro* with PMA and ionomycin.
